# Changes in Autism Traits from Early Childhood to Adolescence in the Study to Explore Early Development

**DOI:** 10.1101/2025.04.16.25325938

**Authors:** Gabriel Dichter, Kyle F. Grosser, Kaitlin K. Cummings, Chyrise Bradley, Tanya P. Garcia, Amanda L Tapia, Rebecca E. Pretzel, Cy Nadler, Eric J. Moody, Brian Barger, Nuri M. Reyes, M. Daniele Fallin, Julie L. Daniels

**Author notes:** All authors declare no conflicts of interest and approve of being listed as an author on this manuscript.

## Abstract

**Purpose:** The objectives of this study were to investigate associations between co-occurring developmental, psychiatric, behavioral, and medical symptoms and conditions and autism spectrum disorder (ASD) traits, as well as predictors of changes in autistic traits from early childhood to adolescence.

**Methods:** Participants from the Study to Explore Early Development (SEED) were identified as having autism spectrum disorder (ASD) (n=707), another developmental disorder (DD) (n=995), or as a population comparison group (POP) (n=898). Caregivers completed the Social Responsiveness Scale-2nd edition (SRS-2) to measure autistic traits and were asked about co-occurring symptoms and conditions when their child was 2-5 years old and 12-16 years old. Children completed the Mullen Scales of Early Learning (MSEL) when they were 2-5 years old.

**Results:** Regression models revealed that in early childhood and adolescence, multiple co-occurring symptoms and conditions were significantly associated with higher SRS-2 scores (e.g., motor, sensory, and sleep problems for children with ASD and DD). Within the ASD and DD groups, but not the POP group, lower MSEL scores at childhood were associated with greater increases in SRS-2 scores between early childhood and adolescence.

**Conclusions:** Findings suggest that motor, sensory, and sleep problems may be important intervention targets for ASD and DD youth with elevated SRS-2 scores and that interventions that target cognitive functioning in childhood may be important to modify trajectories of autistic traits from childhood to adolescence.

## Introduction

Autism spectrum disorder (ASD) is a neurodevelopmental disorder characterized by impaired social communication and restricted, repetitive behaviors (American Psychiatric, 2013) that affects 1 in 36 children in the US (Maenner et al., 2023). Autistic traits during early childhood are dynamic and may change over time (Tunc et al., 2021). Although most autistic children retain their diagnosis (Pierce et al., 2019; Zwaigenbaum et al., 2016), it is not uncommon for the symptom presentations of some autistic children to change substantially during development (Fountain et al., 2023; Kim & Byun, 2018; Ozonoff et al., 2015), for children with similar presentations to follow distinct long-term developmental trajectories (Eigsti et al.), and even for some children to lose or gain an ASD diagnosis (Ozonoff et al., 2018; Ozonoff et al., 2015; Zwaigenbaum et al., 2016). Moreover, some children have a symptom presentation that does not clearly define them as autistic or not autistic (Lord, 2018; Ozonoff et al., 2014), but rather these children occupy a diagnostic transition region -- a region that changes over time between ASD and non-ASD, reflecting a fuzzy boundary between the two categories (Haslam, 2002; Landa et al., 2022; Tunç et al., 2021). Up to one third of early diagnoses of ASD (16-30-months-old) are made with some uncertainty (Klaiman et al., 2022) and up to a third of diagnosed toddlers lose their diagnosis by age seven (Harstad et al., 2023). The extent to which these transitions describe a sizeable portion of autistic children has implications for whether ASD is stable or dynamic, whether traditional measures of ASD diagnoses and symptoms are state or trait measures (Tunç et al., 2021), and highlights the importance of early ASD detection and intervention given the potential for ASD symptom presentations to change during childhood (Ozonoff et al., 2018; Ozonoff et al., 2015).

Between 70% to 95% of autistic children and adolescents have at least one co-occurring psychiatric condition (Gjevik et al., 2011; Harstad et al., 2024; Joshi et al., 2010; Kanne et al., 2009; Lai et al., 2019; Leyfer et al., 2006; Rosen et al., 2018; Simonoff et al., 2008), including anxiety disorders, mood disorders, attention deficit hyperactivity disorder (ADHD), obsessive-compulsive disorder (OCD), and oppositional defiant disorder. Similarly, there are elevated rates of co-occurring medical conditions in autistic cohorts (Al-Beltagi, 2021), including neurogenetic disorders (Devitt et al., 2015; Oxelgren et al., 2017), neurological disorders (Devnani & Hegde, 2015; Lamb et al., 2019; Pacheva et al., 2019), sleep disorders (Devnani & Hegde, 2015; Klukowski et al., 2015; Yang et al., 2018), gastrointestinal disorders (Bresnahan et al., 2015; Fulceri et al., 2016; Prosperi et al., 2019; Wasilewska & Klukowski, 2015), and immune disorders, including autoimmune disorders and allergies (Connery et al., 2018; Jyonouchi, 2010; Zimmerman et al., 2007). A recent analysis of 1418 autistic youth in a large integrated health care delivery system reported increases in prevalence of most health conditions from age 14 to 22 years (Malow et al., 2023). More severe autistic symptoms are associated with an increasing number of co-occurring medical symptoms (Aldinger et al., 2015; Soke et al., 2018; Weber & Gadow, 2017), and high rates of co-occurring behavioral, developmental, psychiatric, and medical conditions are associated with exacerbation of autistic symptoms (de Bruin et al., 2007; Leyfer et al., 2006), diminished impact of ASD treatments (Joshi et al., 2010; McDougle et al., 2003), and worse quality of life and adaptive behavior (Kamio et al., 2013; Kraper et al., 2017).

Despite retrospective studies of predictors of diagnostic stability in individuals with a history of ASD diagnoses (Fein et al., 2013; Moulton et al., 2016), no research to date has prospectively examined the association between diverse co-occurring conditions on autistic traits during development. In this study, during early childhood and during adolescence, we examined cross-sectional associations between behavioral, developmental, psychiatric, and medical conditions and symptoms with autistic traits. Additionally, we examined associations between early childhood co-occurring conditions and changes in autistic traits from early childhood to adolescence. Our analysis considered autistic traits within an ASD group as well as within those with other developmental disabilities and in population controls (Powell et al., 2021). We hypothesized that the presence of co-occurring symptoms and conditions would predict more autistic traits both cross-sectionally and longitudinally across groups, with more pronounced associations within the ASD group.

## Methods

### Participants

The cohort of children examined in this study participated in the Study to Explore Early Development (SEED; https://www.cdc.gov/ncbddd/autism/seed.html). They were identified at 2-5 years old as having ASD, other developmental disabilities (DD group), or as population controls (POP group). SEED has been described in detail elsewhere (Schendel et al., 2012; Wiggins, Levy, et al., 2015); briefly, SEED is a multisite, community-based case–control study of children 30–68 months of age (born September 2003–August 2006) designed to evaluate genetic and environmental factors associated with ASD. A child was eligible if he or she lived from six months of age with a caregiver who was fluent in English (English or Spanish at the Colorado and California sites) and could provide legal consent, and if the child was born and continued to live in the catchment areas of the SEED sites located in California, Colorado, Georgia, Maryland, North Carolina, and Pennsylvania. Institutional review boards at CDC and each site approved the SEED protocol.

Children were recruited from early intervention and special education programs, from healthcare providers serving children with disabilities, and from birth records. Classification into the final study groups was based on a comprehensive in-person evaluation (Wiggins, Reynolds, et al., 2015). Upon enrollment, all children were screened for ASD with the Social Communication Questionnaire (SCQ) (Rutter et al., 2003) and underwent an in-person developmental evaluation using the Mullen Scales of Early Learning (MSEL) (Mullen, 1995). The Autism Diagnostic Observation Schedule (ADOS) (Gotham et al., 2007; Lord et al., 1999) and the Autism Diagnostic Interview-Revised (ADI-R) (Lord et al., 1994) were completed for children with a previous diagnosis of ASD, a SCQ score of 11 or above, or when concerns for ASD were noted during administration of the MSEL. A research classification of ASD was made using an ADOS and ADI-R algorithm: an ASD classification was assigned if a child met ADOS criteria and one of three ADI-R relaxed criteria: (1) met the social domain cutoff and was within two points of the communication domain cutoff, or (2) met the communication domain cutoff and was within two points of the social domain cutoff, or (3) met the social domain cutoff and had at least two points on the behavioral domain. The ADI-R criteria were relaxed since the ADI-R was developed to detect individuals with autistic disorder rather than the broader range of ASD sought in SEED. If a child did not meet research criteria for ASD, they were assigned to the DD group or to the POP group (Wiggins, Reynolds, et al., 2015). Children in the DD group were those who were recruited from an education or clinic source and completed a limited developmental evaluation because ASD risk was not indicated on the SCQ, or they completed the comprehensive developmental evaluation but did not meet study criteria for a child with ASD. Children in the POP group were those who were recruited from state vital records and completed a limited developmental evaluation because ASD risk was not indicated on the SCQ, or they completed the comprehensive developmental evaluation but did not meet study criteria for a child with ASD.

This analysis included families who participated in a follow-up survey of parents and guardians of adolescents 12–16 years old who previously participated in SEED. Eligibility for this follow-up study included having a classification of ASD, DD, or POP at 2-5 years old (i.e. those with Incomplete Classification were not eligible for the follow-up study), consenting to being contacted about future studies related to SEED, formerly participating through the Georgia, Maryland, North Carolina, or Pennsylvania sites, and having a caregiver knowledgeable about the child’s health and development. Children defined as Incomplete Classification were those who were asked to complete a comprehensive developmental evaluation but did not complete the evaluation for any reason or completed the evaluation, had a mental age less than 24 months, and the study clinician did not define the child as ASD; these children were excluded from analyses (31% of the total childhood sample).

### Measures

#### Validated Instruments

*Collected at the childhood and adolescent timepoints from caregivers:*

##### Social Responsiveness Scale-2nd edition (SRS-2)

The magnitude of autistic traits was assessed using parent report on the Social Responsiveness Scale-2nd edition (SRS-2) (Constantino & Gruber, 2012). This 65-item rating scale measures deficits in social behavior associated with ASD as characterized in the 4^th^ edition of the Diagnostic and Statistical Manual of Mental Disorders. Scores <60 are considered not clinically significant. The mild range includes scores of 60 to 65, which indicate mild to moderate deficiencies in social behavior. Scores that fall between 66 and 75 are considered moderate, signaling some clinically significant social deficits; scores of 76 or higher are considered severe, suggesting clinically significant deficits in social functioning that interfere with interactions with others (Constantino & Gruber, 2012; Moody et al., 2017). All analyses used SRS-2 total t-scores (mean=50, SD=10). SRS-2 scores were the primary outcome measure in all cross-sectional and longitudinal analyses.

##### The Children’s Sleep Habits Questionnaire (CSHQ)

The CSHQ is a 33-item parent-report sleep screener designed for school-aged children that yields a total sleep disturbance score with adequate internal consistency and reliability for community and clinical samples (Owens et al., 2000; Reynolds et al., 2019). The CSHQ reflects key sleep domains that encompass the major medical and behavioral sleep disorders in this age group: bedtime resistance, sleep duration, parasomnia, sleep disordered breathing, night wakings, daytime sleepiness, sleep anxiety, and sleep onset delay (Owens et al., 2000). The total sleep disturbance score was used to measure childhood associations; the sleep duration and parasomnia subscales were used to measure adolescent associations because these were the only subscales administered at that timepoint. Higher CSHQ scores indicate more sleep problems.

##### Caregiver-Reported Conditions

Caregivers answered questions about their child’s diagnosed health conditions using a yes/no response pattern. <u>At the childhood timepoint</u>, caregivers were asked if their child had ever received a diagnosis of birth defect, pneumonia, and respiratory syncytial virus. At the childhood and adolescent timepoints, caregivers were asked if their child had ever received a diagnosis of allergy/eczema, asthma, epilepsy or seizures, gastrointestinal symptoms (GIS) (e.g., diarrhea, constipation), hearing problems, movement or motor problems, and several behavioral, developmental or psychiatric conditions (i.e., ADHD, learning disability, self-injurious behavior, sensory integration disorder, and speech or language disorder). <u>At the</u> <u>adolescent timepoint</u>, caregivers were asked if their child had ever received a diagnosis of anxiety, behavioral or conduct problems, depression, or dyslexia.

For the adolescent timepoint, a learning disability variable was derived by combining questions about having a learning disability and dyslexia. An internalizing variable was derived by combining questions about anxiety and depression and an externalizing variable was derived by combining questions about ADHD, ODD, behavioral or conduct problems (note that these derived variables represent internalizing and externalizing conditions, whereas the CBCL subscales index internalizing and externalizing symptoms).

*Collected at the childhood timepoint only*:

##### Mullen Scales of Early Learning

Administered to children, the MSEL is a standardized developmental assessment for infants and preschool-age children up to 68 months of age. It is divided into five subscales: (1) gross and (2) fine motor skills, (3) receptive and (4) expressive language, and (5) visual reception (Mullen, 1995). The last four subscales can be combined to create an overall Early Learning Composite (ELC) standard score (mean=100, SD=15), a measure of fluid intelligence thought to underlie general cognitive ability. The MSEL ELC demonstrates high internal consistency and convergent validity with other measures of cognitive ability among children with and without ASD (Bishop et al., 2011). Higher MSEL standard scores indicate more advanced skill development.

##### Child Behavior Checklist (CBCL)

Reported by caregivers, the CBCL is a standardized questionnaire for infants and preschool-age children from 18 months to 68 months of age. The CBCL assesses behavioral, emotional, and social over the past two months (Achenbach & Rescorla, 2000). The CBCL includes 99 items across empirically based syndromic scales, some of which are grouped into broad-spectrum externalizing (i.e., attention problems and aggressive behavior) and internalizing (e.g., anxious/depressed and withdrawn) problem scales. CBCL externalizing and internalizing t-scores >59 indicate borderline to clinically significant problems. Higer CBCL t-scores indicate more problem behaviors.

### Analytic Plan

In preliminary analyses, we summarized characteristics of the samples by study group and assessed distributions of diagnosed conditions and SRS-2 t-scores at childhood and adolescence. Between group differences in diagnosed conditions were assessed with chi-square analyses.

Changes in SRS-2 scores from childhood to adolescence were assessed for each of the diagnostic groups using paired t-tests. To limit the number of analyses conducted, predictors were selected for analysis if they showed greater than 5% prevalence in the ASD sample at childhood, with the exception of epilepsy or seizures which were endorsed by only 3.5% of the ASD sample at childhood, but was evaluated due to evidence of shared pathophysiology of epilepsy and ASD (Frye et al., 2016) and elevated rates of seizures for autistic individuals (Canitano, 2007).

For childhood cross-sectional analyses, separate linear regression models for each study group evaluated the associations between childhood SRS-2 scores and the following predictors: the MSEL ELC standard score, CBCL externalizing t-score, CBCL internalizing t-score, the CSHQ total score; and conditions from the caregiver-reported questionnaire: allergies/eczema, ADHD, asthma, birth defects, epilepsy or seizures, GIS, hearing problems, learning disabilities, motor or movement problems, pneumonia, respiratory syncytial virus, self-injurious behavior, sensory integration disorder, and speech or language disorder.

For adolescent cross-sectional analyses, separate linear regression models for each study group evaluated the associations between adolescent SRS-2 scores and the following predictors: CSHQ sleep duration and parasomnia scores, and conditions from the caregiver-reported questionnaire: allergies/eczema, ADHD, anxiety, asthma, behavioral or conduct problems, depression, developmental concerns, epilepsy or seizures, GIS, learning disabilities, motor or movement problems, sensory integration disorder, self-injurious behaviors, and speech or language disorder.

For longitudinal analyses, linear regression models evaluated associations between co-occurring conditions and symptoms and changes in SRS-2 scores from childhood to adolescence. Each model estimated the association for one predictor at a time. Analyses of change in SRS-2 scores included participants with SRS-2 data available at childhood and adolescent timepoints. All regression models were adjusted for study site (CA, CO, GA, MD, NC, PA), child sex (male, female), child age, and maternal demographic factors (age, education (<= some college, >= bachelor’s degree), and percent poverty level (<=300%, >300%)); longitudinal analyses also included SRS-2 scores at childhood as a covariate. Beta values, 95% confidence intervals and *p*-values are reported.

All continuous covariates and predictors were centered and scaled across diagnostic groups (separately for the childhood and adolescent datasets) by subtracting their sample means and then dividing by their sample standard deviations to reduce possible collinearity. Potential interactions between predictors and diagnostic group were assessed using interaction terms. Associations between each predictor and SRS-2 score were then assessed within the ASD, DD, and POP groups separately. The *p*-values for associations of each predictor were corrected for a false discovery rate of 0.05 (Murray & Blume, 2021) within analyses for each timepoint separately. Missing data were addressed by removing incomplete cases on a “per-model” basis. All analyses were done in R (Team, 2013).

## Results

At the childhood timepoint, 898 children were included in the POP group, 707 children were included in the ASD group, and 995 children were included in the DD group. At the adolescent timepoint, there were 313 adolescents in the POP group, 208 adolescents in the ASD group, and 345 adolescents in the DD group. At both timepoints, diagnostic groups differed on the following sociodemographic variables: sex distribution (reflecting the greater proportion of males in the ASD group than in the other two groups and a greater proportion of males in the DD group than in the POP group), maternal education (reflecting a larger proportion attending college in the POP group than the other two groups), percent poverty (reflecting a larger proportion greater than 300% of the poverty rate in the POP group than the other two groups), and maternal race (Table 1).

**Table 1.**
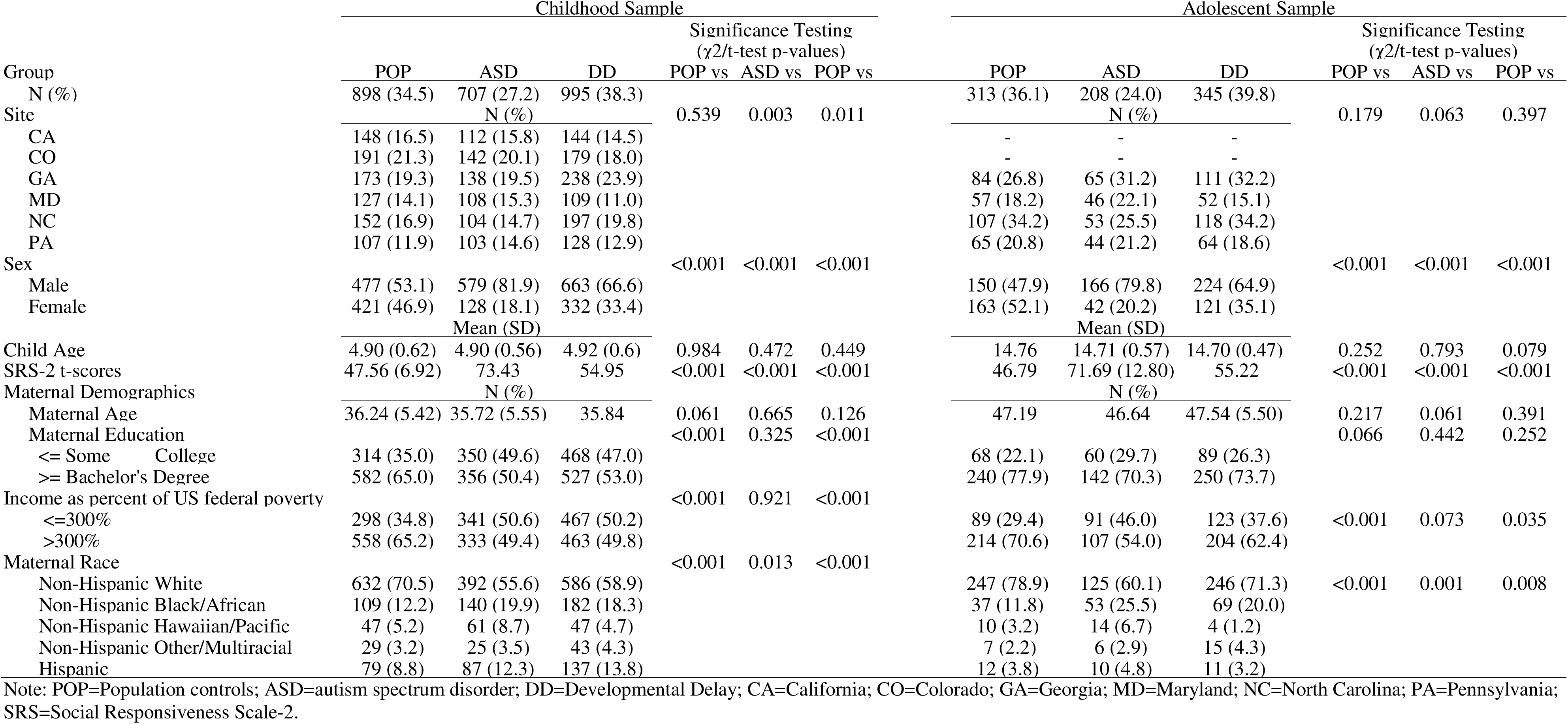
Characteristic of participants enrolled in the Study to Explore Early Development – Phase 1 (2007-2011; left) and SEED Teen (2017-2021; right) data.

Compared to participants without adolescent data, participants in the adolescent sample were younger at the childhood timepoint (*p*<.01), had a higher proportion of female participants (*p*<.05), had a greater proportion whose mothers had at least a bachelor’s degree (*p*<.001), had a greater proportion at least 300% above the poverty rate (*p*<.001), had a greater proportion whose mothers identified as white (*p*<.001), and had lower SRS-2 scores (*p*<.001). The distributions of childhood and adolescent predictors, and whether these differed between groups, are presented in *Supplemental Tables 1 and 2*)

### SRS-2 score distributions and changes

The mean SRS-2 score for the ASD group was above the suggested clinical cutoff of 60 (mean=73.43) and the mean SRS-2 score2 for the DD and POP groups were below the suggested clinical cutoff of 60 (means=54.95 and 47.56, respectively), reflecting the dimensional nature of SRS-2 scores. Figure 1 illustrates mean SRS-2 scores and changes in SRS-2 scores from childhood to adolescence in the ASD (t(190)=1.36, *p*=0.18), DD (t(320)=-2.55, *p*=0.01), and POP groups (t(298)=-0.25, *p*=0.80), indicating significant change over time in the DD group only.

**Figure 1.**
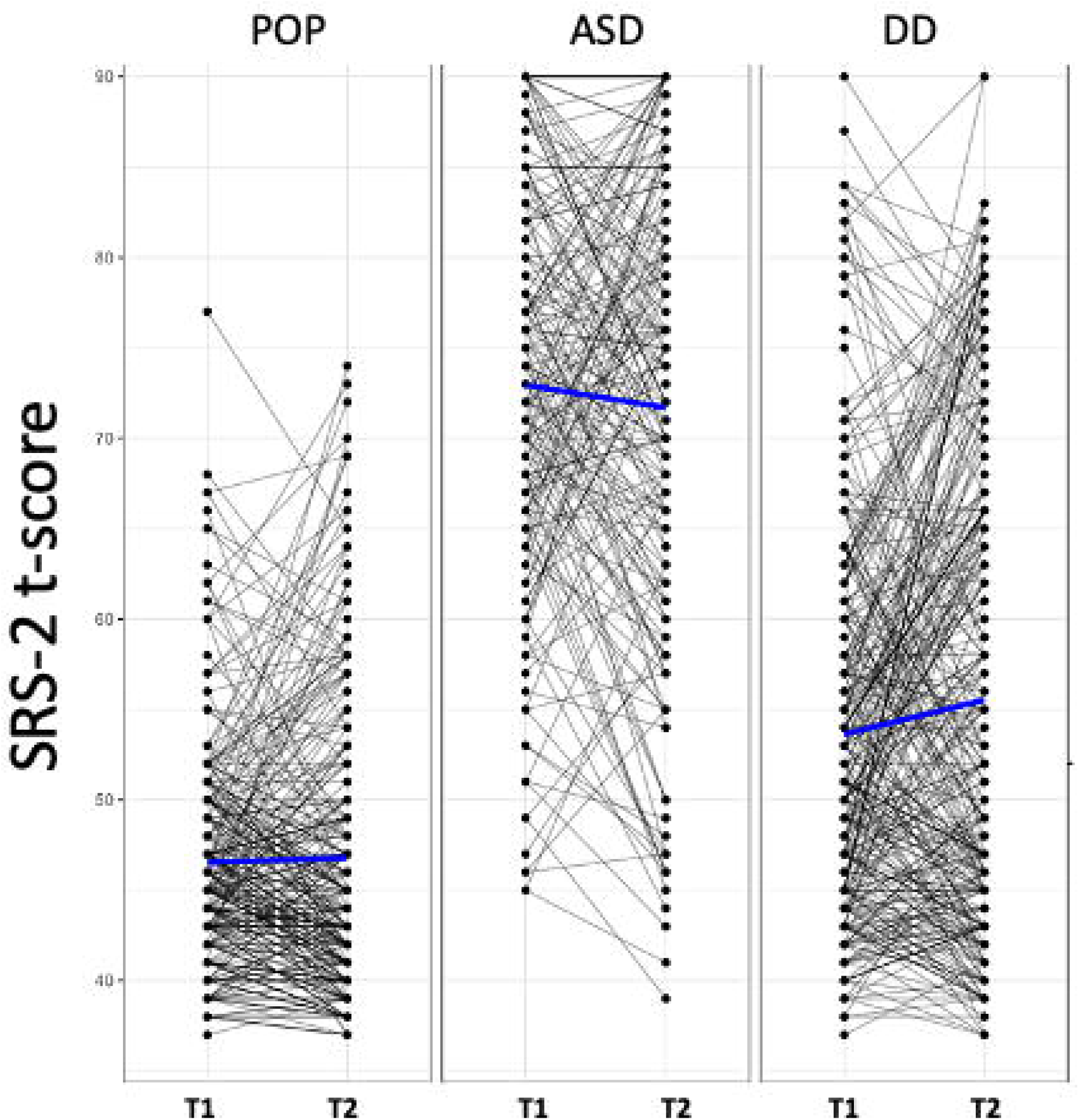
Change in SRS-2 scores from Childhood to Adolescence by Group Figure 1. Spaghetti plots of SRS-2 score trajectories from childhood to adolescence in the population controls (left), ASD (middle), and developmental delay (right) groups. Dots reflect SRS-2 scores and may thus reflect more than one participant. Black lines indicate participant-level data. Blue lines indicate group means. Note: POP=population Controls; ASD=autism spectrum disorder; DD: developmental delay; SRS=Social Responsiveness Scale. T1: Childhood (ages 2-5; 2007-2011). T2: Adolescent (ages 12-16; 2017-2021).

### Childhood cross-sectional predictors of SRS-2 scores

Childhood cross-sectional predictors of childhood SRS-2 scores are presented in Figures 2 and 3 (parameter estimates and p-values associated with these figures are provided in *Supplemental Table 3*). Among validated instruments, cognitive functioning measured with the MSEL ELC standard score, CBCL internalizing t-score, CBCL externalizing t-score, and CSHQ total sleep score were associated with significantly higher SRS-2 scores in all three study groups. Among diagnosed conditions, ADHD, GIS, and sensory integration disorder were associated with significantly higher SRS-2 scores in all three study groups. Self-injurious behaviors were associated with significantly higher SRS-2 scores within the ASD and DD groups; an estimate for self-injurious behaviors in the POP group is not provided due to small sample size. Other variables associated with higher SRS-2 scores based on study group were allergies/eczema (POP), epilepsy/seizures (ASD), hearing problems (POP), learning disability (DD and POP), movement or motor problems (DD and POP), and speech or language disorder (DD and POP).

**Figure 2.**
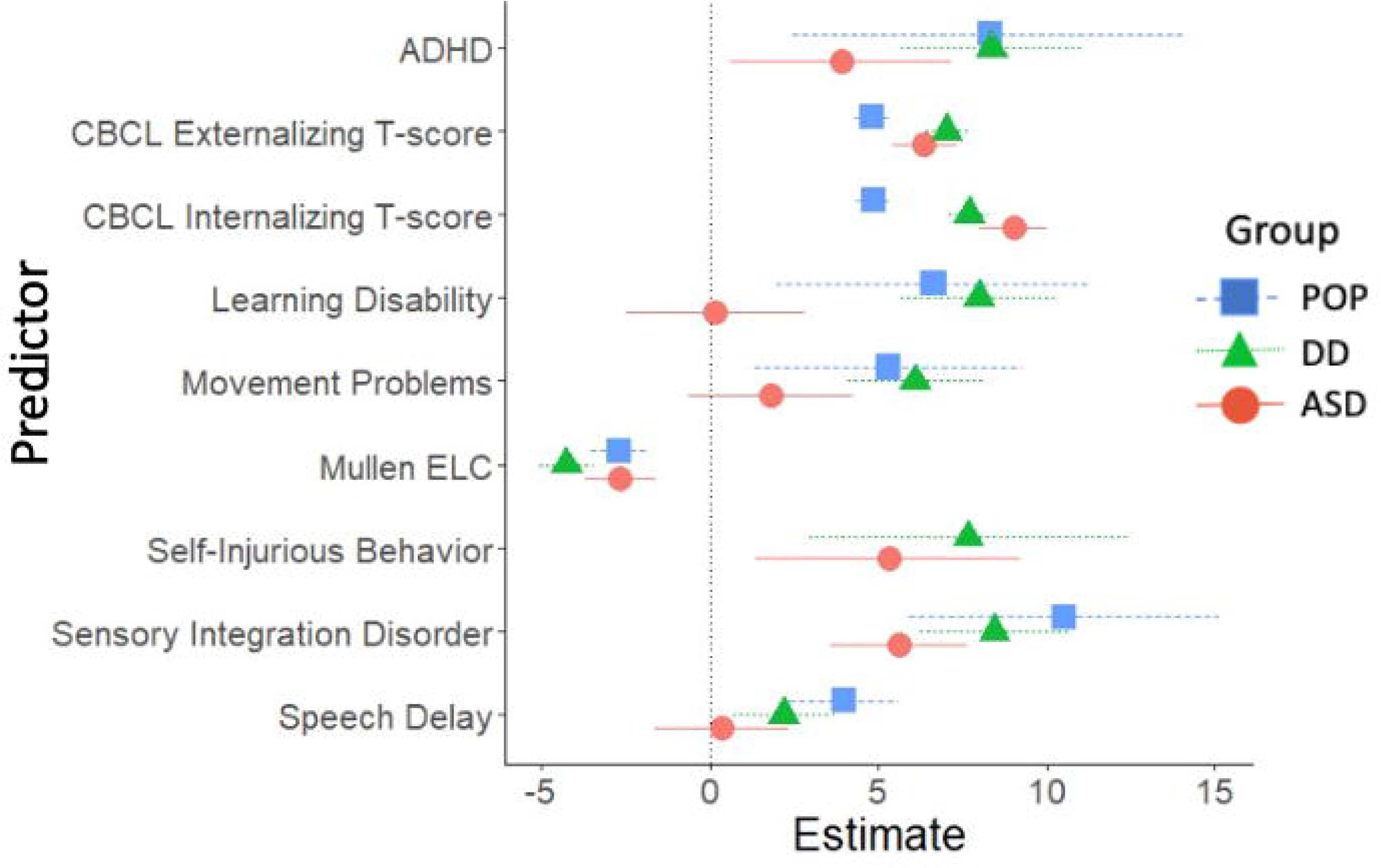
Cross-Sectional Developmental, Behavioral, and Psychiatric Predictors of SRS-2 Scores in Childhood (ages 2-5 years), Study to Explore Early Development – Phase 1 (2007-2011). Figure 2. Forrest plots depicting relations between developmental, behavioral, and psychiatric predictors of SRS-2 scores in childhood. Horizontal lines indicate the 95% confidence interval surrounding the point estimate of the effect of each predictor on SRS-2 scores at childhood. The plot depicts model results for the POP, ASD, and DD groups separately. Models controlled for study site, child sex, child age at the Mullen assessment, maternal age, maternal education, and maternal percent poverty level. The x-axis reflects regression coefficients from the models evaluated. Point estimates reflect change in SRS-2 score per 1 SD change in the predictor (for continuous predictors) or given the presence of the predictor (for binary predictors). Note: ADHD=attention deficit hyperactivity disorder; ASD=autism spectrum disorder; POP=population controls; DD=developmental delay; ELC=Early Learning Composite; SRS=Social Responsiveness Scale.

**Figure 3.**
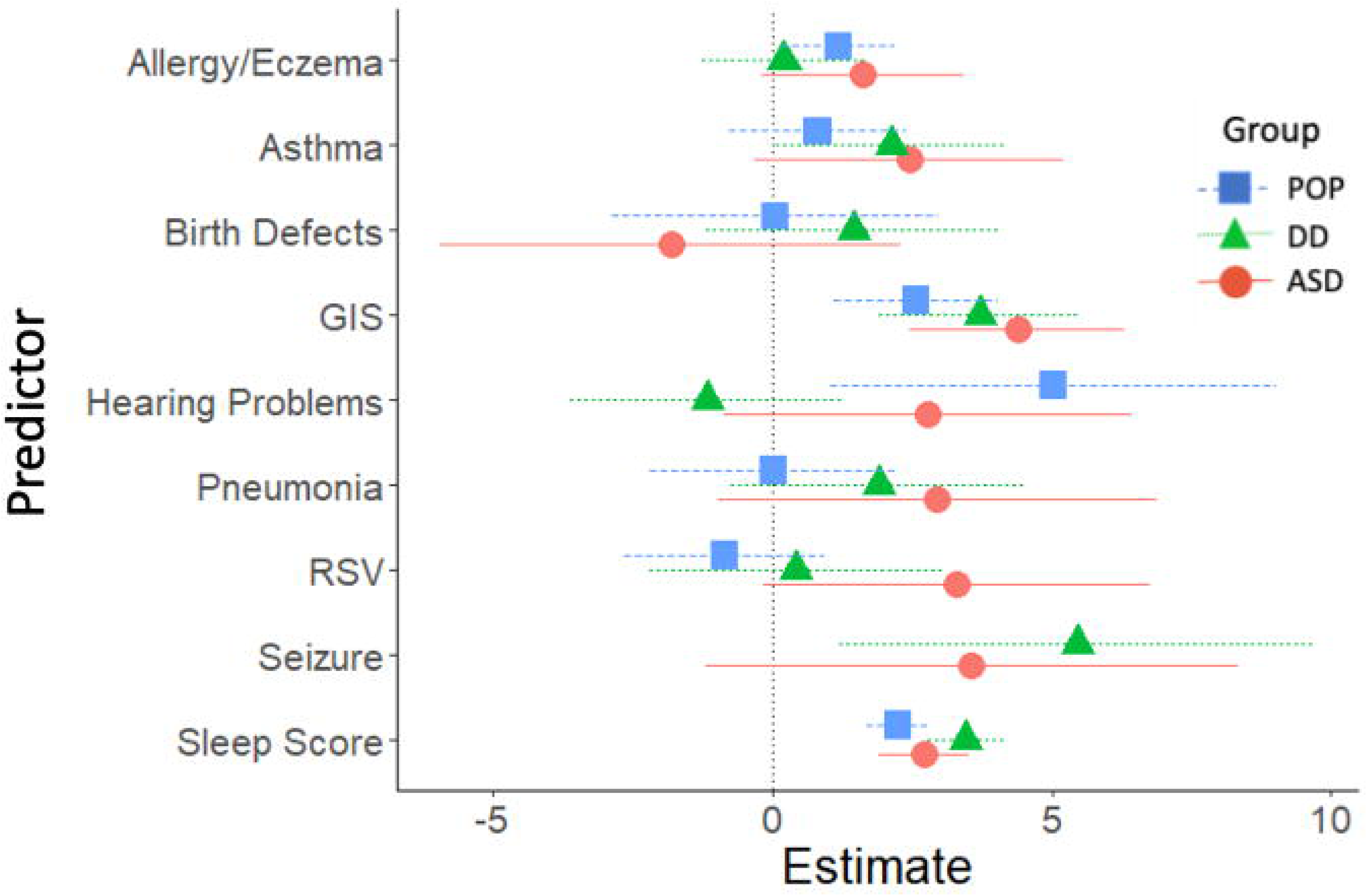
Other Cross-Sectional Predictors of SRS-2 Scores in Childhood (ages 2-5 years; 2007-2011), Study to Explore Early Development – Teen (2017-2021). Figure 3. Forrest plots depicting relations between other predictors of SRS-2 scores in childhood. Horizontal lines indicate the 95% confidence interval surrounding the point estimate of the effect of each predictor on SRS-2 scores at childhood. The plot depicts model results for the POP, ASD, and DD groups separately. Models controlled for study site, child sex, child age at the Mullen assessment, maternal age, maternal education, and maternal percent poverty level. Models for the combined sample did not include case status as an interaction term. To aide with figure scaling, seizure and self-injury are not depicted for the POP group and seizures are not depicted in the total sample due to low prevalence. The x-axis reflects regression coefficients from the models evaluated. Point estimates reflect change in SRS-2 score per 1 SD change in the predictor (for continuous predictors) or given the presence of the predictor (for binary predictors). Note: ASD=autism spectrum disorder; POP=population controls; DD=developmental delay; SRS=Social Responsiveness Scale GIS=gastrointestinal symptoms.

### Adolescent cross-sectional predictors of SRS-2 scores

Adolescent cross-sectional predictors of adolescent SRS-2 scores are presented in Figures 4 and 5 (parameter estimates and p-values associated with these figures are provided in *Supplemental Table 4*). Among validated instruments, CSHQ sleep duration was associated with significantly higher SRS-2 scores in all three study groups. Among diagnosed conditions, ADHD, behavior or conduct problems, learning disability, and speech or language disorder were associated with significantly higher SRS-2 scores in all three study groups. Other variables associated with higher SRS-2 scores based on study group were anxiety (DD and POP), depression (DD and POP), GIS (ASD and DD), movement or motor problems (ASD and DD), parasomnia (ASD and DD), self-injurious behaviors (ASD and DD) and sensory integration disorder (ASD and DD).

**Figure 4.**
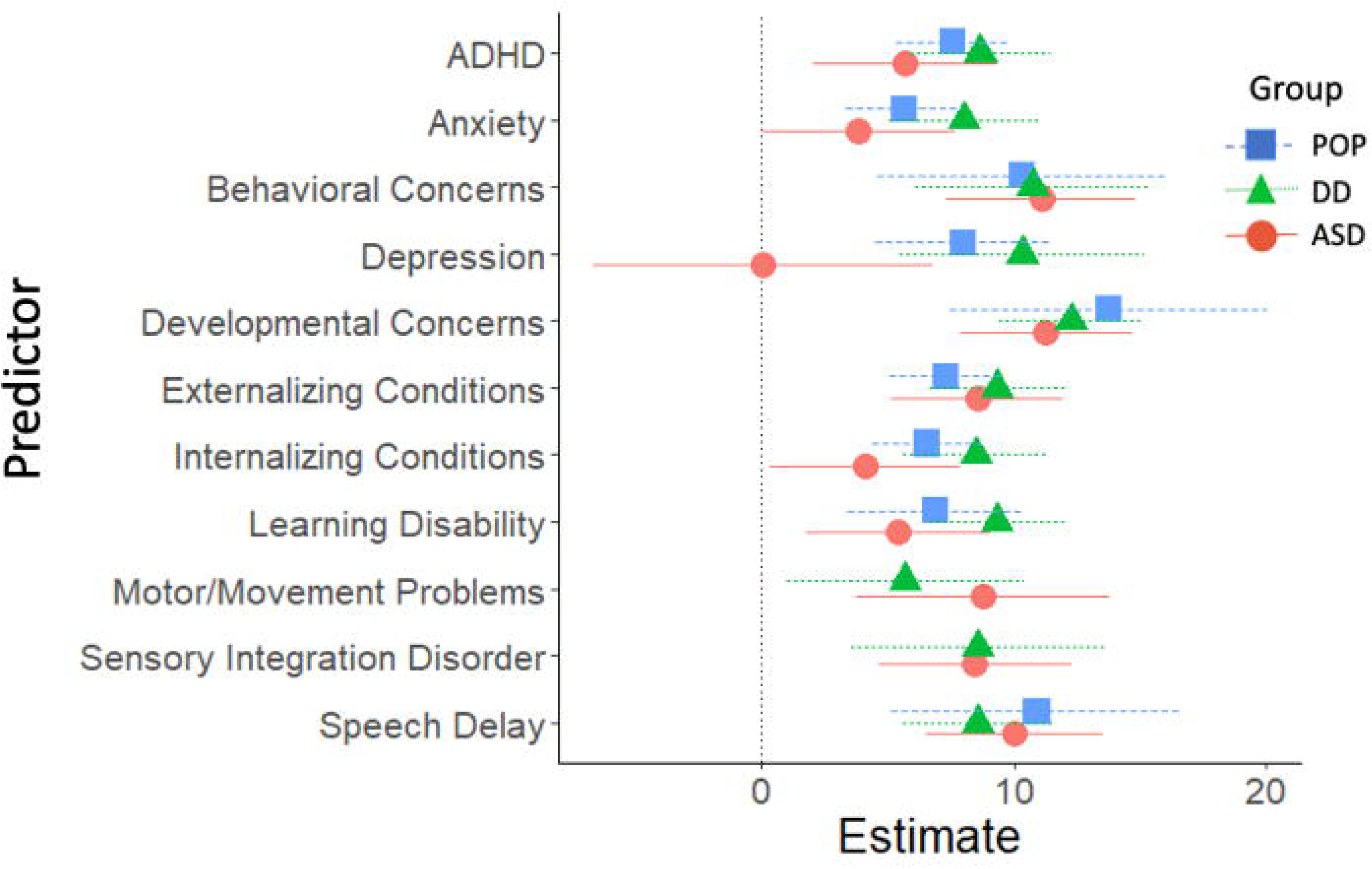
Cross-Sectional Developmental, Behavioral, and Psychiatric Predictors of SRS-2 Scores in Adolescence (ages 12-16 years), Study to Explore Early Development – Teen (2017-2021). Figure 4. Forrest plots depicting relations between developmental, behavioral, and psychiatric predictors of SRS-2 scores in adolescence. Horizontal lines indicate the 95% confidence interval surrounding the point estimate of the effect of each predictor on SRS-2 scores at adolescence. The plot depicts model results for the POP, ASD, and DD groups separately. Models controlled for study site, child sex, child age at the Mullen assessment, maternal age, maternal education, and maternal percent poverty level. The x-axis reflects regression coefficients from the models evaluated. Point estimates reflect change in SRS-2 score per 1 SD change in the predictor (for continuous predictors) or given the presence of the predictor (for binary predictors). Note: ADHD=attention deficit hyperactivity disorder; ASD=autism spectrum disorder; POP=population controls; DD=developmental delay; SRS=Social Responsiveness Scale.

**Figure 5.**
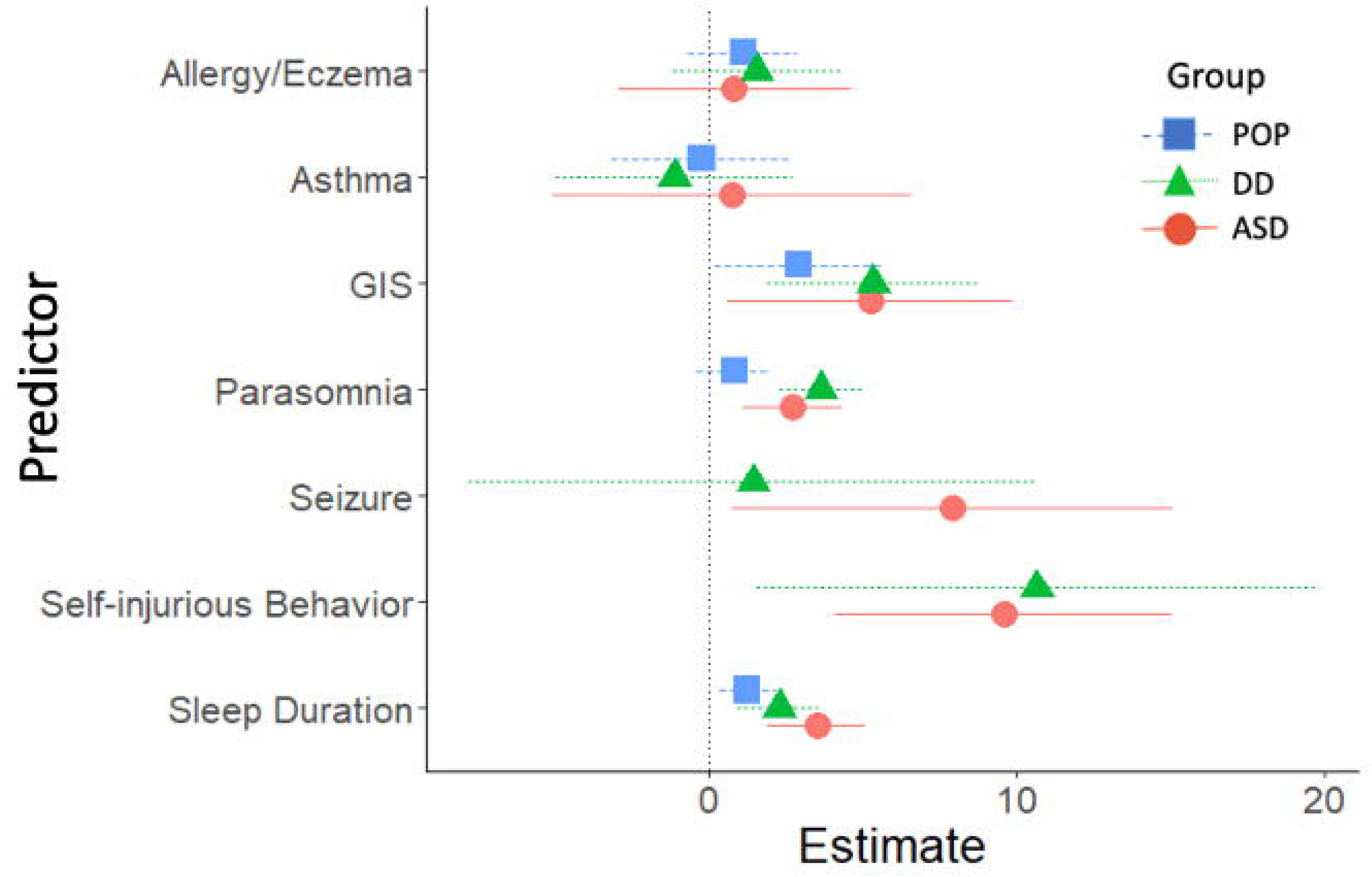
Other Cross-Sectional Predictors of SRS-2 Scores in Adolescence (ages 12-16 years), Study to Explore Early Development – Teen (2017-2021). Figure 5. Forrest plots depicting relations between other predictors of SRS-2 scores in adolescence. Horizontal lines indicate the 95% confidence interval surrounding the point estimate of the effect of each predictor on SRS-2 scores at adolescence. The plot depicts model results for the POP, ASD, and DD groups separately. Models controlled for study site, child sex, child age at the Mullen assessment, maternal age, maternal education, and maternal percent poverty level. To aide with figure scaling, seizure, self-injurious behaviors, sensory integration disorder, and motor problems are not depicted in the POP group due to low prevalence. The x-axis reflects regression coefficients from the models evaluated. Point estimates reflect change in SRS-2 score per 1 SD change in the predictor (for continuous predictors) or given the presence of the predictor (for binary predictors). Note: ASD=autism spectrum disorder; POP=population controls; DD=developmental delay; SRS=Social Responsiveness Scale.

### Longitudinal analyses: childhood predictors of SRS-2 change scores

Longitudinal predictors of change in SRS-2 scores from childhood variables are presented in Figures 6 and 7 (parameter estimates and p-values associated with these figures are provided in *Supplemental Table 5*). Longitudinal analyses revealed that in the ASD and DD groups, lower MSEL ELC standard scores in childhood were associated with greater increases in SRS-2 scores between the childhood and adolescence timepoints (Figure 8). In the POP group alone, learning disability was associated with greater increases in SRS-2 scores between the childhood and adolescence timepoints and in the DD group alone, self-injurious behaviors were associated with greater increases in SRS-2 scores between the childhood and adolescence timepoints.

**Figure 6.**
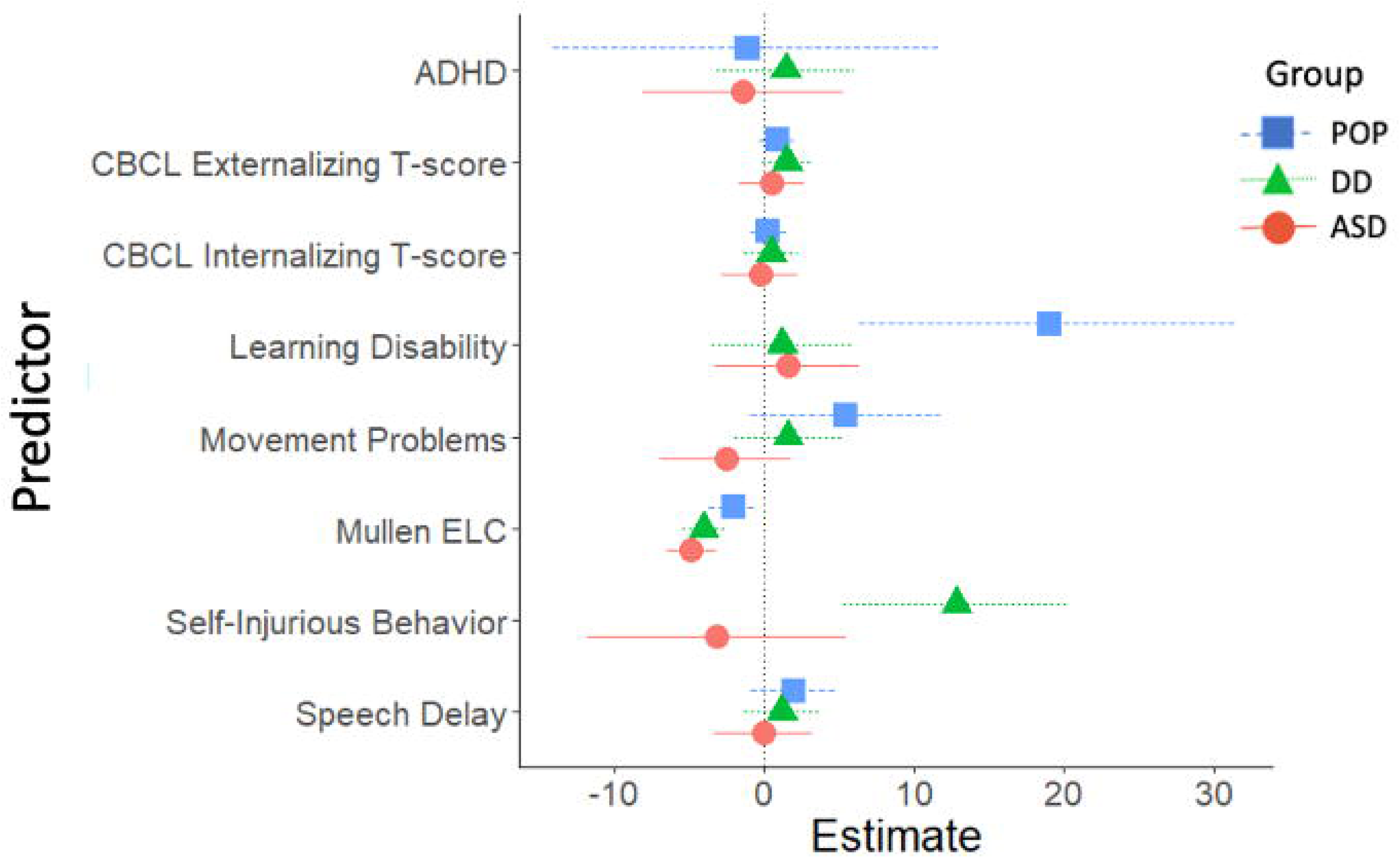
Developmental, Behavioral, and Psychiatric Predictors of Change in SRS-2 from Childhood to Adolescence (ages 12-16 years), Study to Explore Early Development – Teen (2017-2021). Figure 6. Forrest plots depicting relations between developmental, behavioral, and psychiatric predictors of SRS-2 change scores from childhood to adolescence. Horizontal lines indicate the 95% confidence interval surrounding the point estimate of the effect of each predictor on SRS-2 change scores. The plot depicts model results for the POP, ASD, and DD groups separately. Models controlled for study site, child sex, child age at the Mullen assessment, maternal age, maternal education, and maternal percent poverty level. Point estimates reflect change in SRS-2 score per 1 SD change in the predictor (for continuous predictors) or given the presence of the predictor (for binary predictors). Note: ADHD=attention deficit hyperactivity disorder; ASD=autism spectrum disorder; POP=population controls; DD=developmental delay; ELC=Early Learning Composite; SRS=Social Responsiveness Scale.

**Figure 7.**
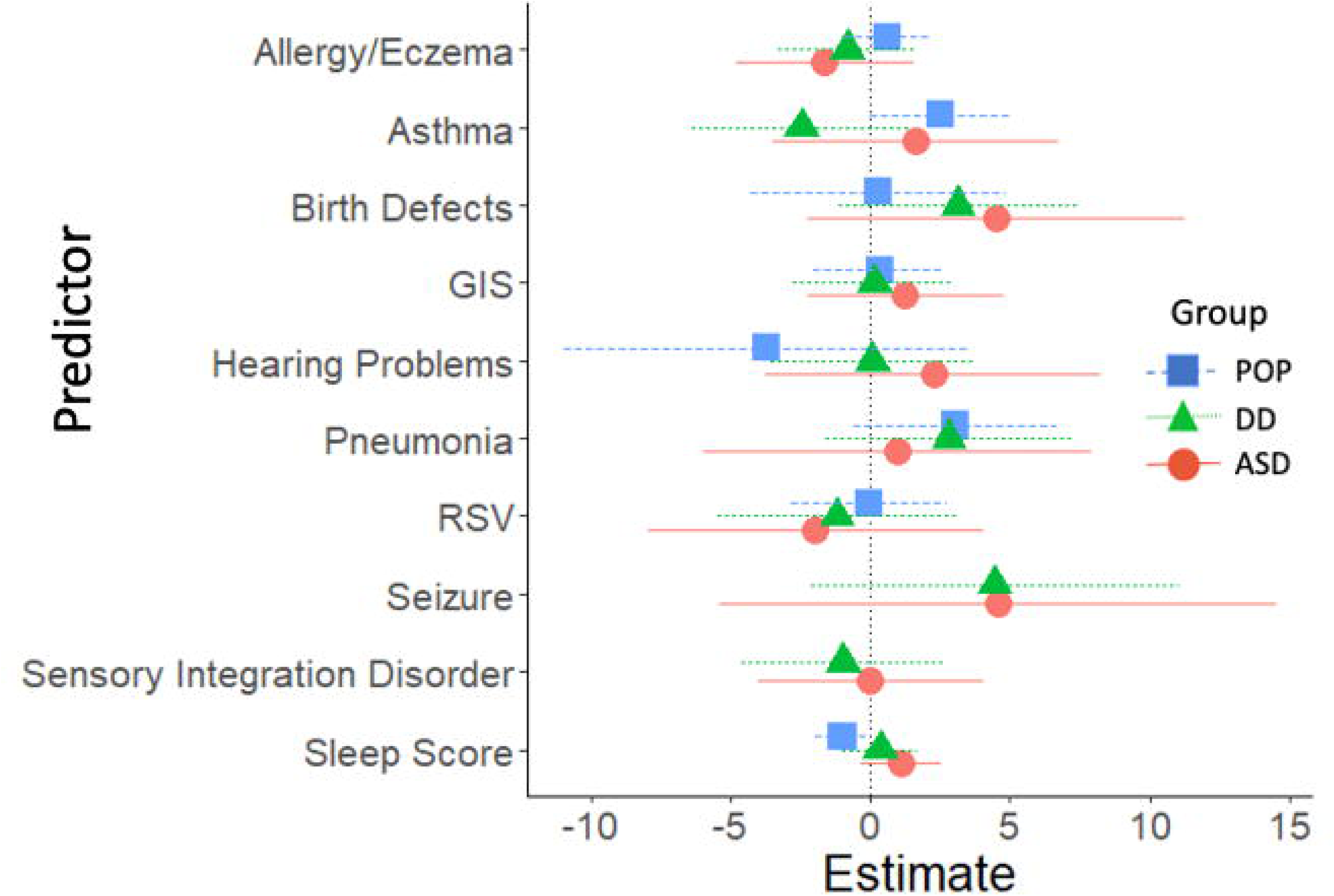
Other Predictors of Change in SRS-2 from Childhood to Adolescence (ages 12-16 years), Study to Explore Early Development – Teen (2017-2021). Figure 7. Forrest plots depicting relations between other predictors of SRS-2 change scores from childhood to adolescence. Horizontal lines indicate the 95% confidence interval surrounding the point estimate of the standardized effect of each predictor on SRS-2 change scores. The plot depicts model results for the POP, ASD, and DD groups separately. Models controlled for study site, child sex, child age at the Mullen assessment, maternal age, maternal education, and maternal percent poverty level. To aid with figure scaling, seizure and self-injury are not depicted for the POP group due to low prevalence. Point estimates reflect change in SRS-2 score per 1 SD change in the predictor (for continuous predictors) or given the presence of the predictor (for binary predictors). Note: ASD=autism spectrum disorder; POP=population controls; DD=developmental delay; SRS=Social Responsiveness Scale; GIS=gastrointestinal symptoms.

**Figure 8.**
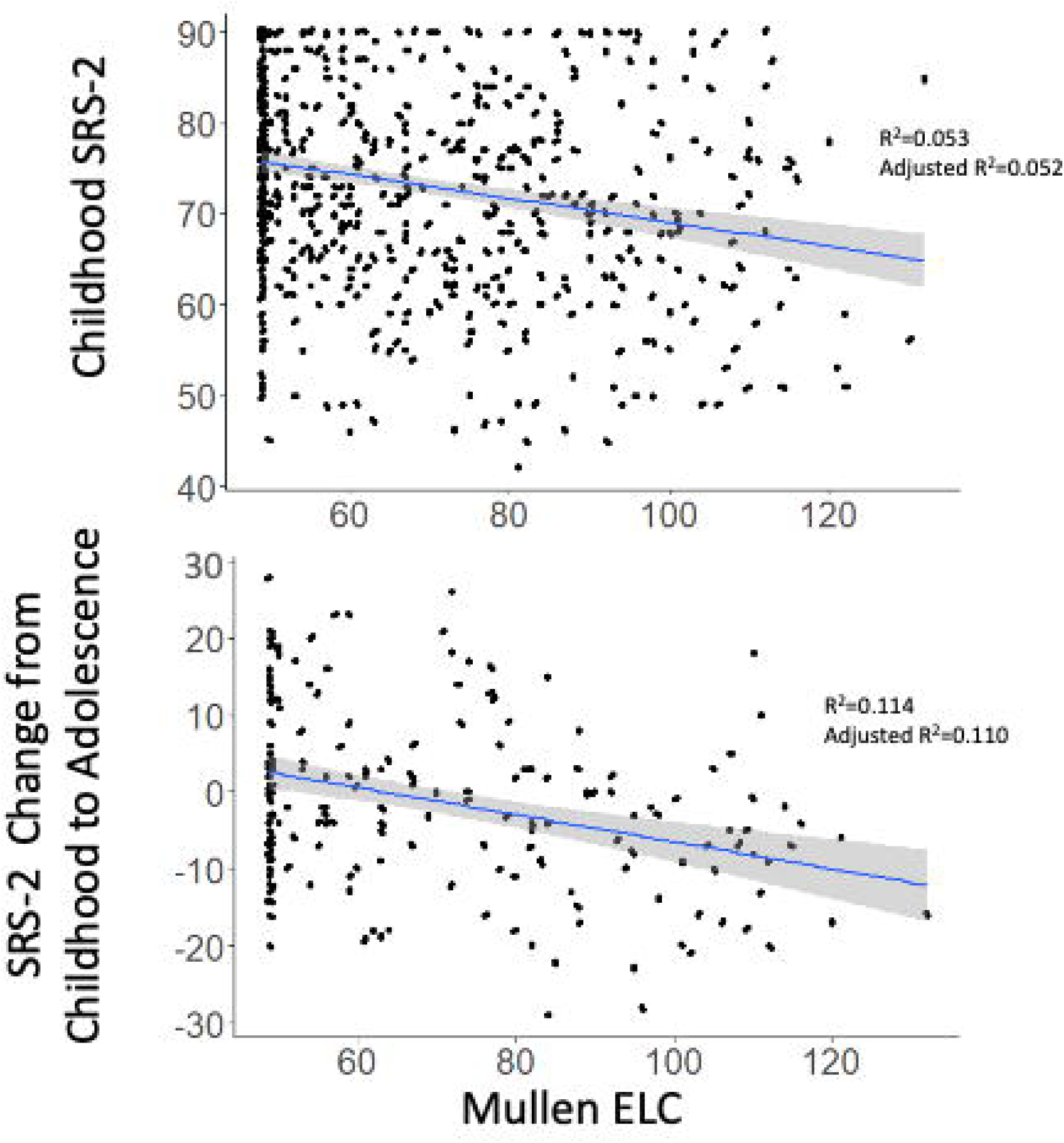
Scatterplots depicting relations between SRS-2 and Mullen ELC scores in the total sample and in the ASD group. Figure 8. Scatterplots depicting relations between childhood Mullen ELC and childhood SRS-2 (top panel) and SRS-2 change between childhood to adolescence (bottom panel) for the ASD group. Note: ASD=autism spectrum disorder; POP=population controls; DD=developmental delay; ELC=Early Learning Composite; SRS=Social Responsiveness Scale.

## Discussion

We examined the influence of co-occurring developmental, behavioral, psychiatric, and medical symptoms or conditions on autistic traits in children with and without ASD in childhood, in adolescence, and on change in autistic traits between these two developmental periods in the SEED sample. Results lend further support to prior findings that co-occurring conditions are common in ASD. Cross-sectional analyses revealed that ADHD and sleep problems were associated with SRS-2 scores in all three study groups at both timepoints. Additionally, GIS, self-injurious behaviors, and sensory integration disorder were associated with SRS-2 scores at both timepoints in the ASD and DD groups; CBCL externalizing scores, CBCL internalizing scores, epilepsy/seizure disorder, and MSEL ELC scores were associated with SRS-2 scores in childhood in the ASD group; and movement or motor disorders were associated with SRS-2 scores in adolescence in the ASD and DD groups. MSEL ELC scores were associated with longitudinal increases in SRS-2 scores in the ASD and DD groups and self-injurious behaviors were associated with longitudinal increases in SRS-2 scores in the DD group.

Across all cross-sectional analyses, significant associations were detected in the hypothesized direction such that greater intensity or frequency of a condition, symptom, or disorder was associated with greater SRS-2 scores. This is consistent with findings that autistic children and adolescents experience higher prevalence of co-occurring psychiatric conditions (Gjevik et al., 2011; Joshi et al., 2010; Kanne et al., 2009; Khachadourian et al., 2023; Lai et al., 2019; Leyfer et al., 2006; Rosen et al., 2018; Simonoff et al., 2008) which, in turn, increases the risk of poorer long-term outcomes (Bryson et al., 2008; Close et al., 2012; Su et al., 2022) including higher mortality (Cummings et al., 2016). Additionally, in the first three years of life, increased rates of neurological, nutrition, genetic, ear, nose, and throat, and sleep conditions are associated with an increased likelihood of subsequent ASD diagnoses (Alexeeff et al., 2017). Moreover, the rates of most health conditions increase in autistic individuals from age 14 to 22 years (Malow et al., 2023), and co-occurring psychiatric conditions with ASD are known to persist from childhood into adolescence (Joshi et al., 2010).

It is noteworthy that many of the predictors of autistic traits were evident in the control and DD groups as well as in the ASD group. The characterizing features of ASD are continuously distributed in the population (Constantino, 2011; Wagner et al., 2019), and these findings are consistent with the potential for health and psychiatric factors to canalize the development of biological systems that contribute to the emergence of autistic traits in those below as well as those above a cutoff for an ASD diagnosis (Shultz et al., 2018). Indeed, a recent study of autistic traits in monozygotic twins demonstrated that the pursuit for autism biomarkers of genetic risk for ASD should also include individuals below the clinical threshold for diagnosis (Castelbaum et al., 2020) because individuals without an ASD diagnosis may carry sub-threshold traits that exert causal influences on the development of ASD (Constantino, 2021). It is also notable that sensory and sleep challenges were significantly associated with SRS-2 scores for children with ASD and DD at both developmental timepoints, suggesting that interventions that target these symptoms may influence the expression of ASD traits. It is possible that nonspecific causal factors may influence the development of both ASD and co-occurring conditions (Hawks & Constantino, 2020).

The multiple significant cross-sectional associations between SRS-2 scores and co-occurring conditions and symptoms provide clues for intervention targets that may reduce autistic traits, however cross-sectional associations do not necessarily imply causal relations. The only significant childhood predictor of changes in autistic traits from longitudinal analyses was the association between MSEL-ELC scores at early childhood and change in autistic traits from early childhood to adolescence within the ASD and DD groups. For every standard deviation increase in MSEL-ELC scores, SRS-2 scores decreased by 3.92 points. This association is consistent with multiple reports of relations between cognitive functioning and social functioning in autistic children (Isaksson et al., 2019; Morrison et al., 2019; Peterson et al., 2015; Sasson et al., 2020), an association that is not commonly present in typically-developing samples (August et al., 2012; Mohn et al., 2014; Peterson et al., 2015). Additionally, cognitive functioning has been shown to differentiate 12- and 24-month-olds at high probability of developing ASD from low probability counterparts (Estes et al., 2015), corroborating that cognitive functioning during very early childhood may be predictive of later autistic traits.

This longitudinal relation between cognitive functioning and changes in autistic traits suggests an important public health implication of this research, namely that interventions impacting cognitive functioning in autistic children may result in improved social communication later in development for families who seek treatment. Cognitive functioning is a primary target of some early intensive interventions for ASD designed to improve social communication, including the Early Start Denver Model (ESDM; Dawson et al., 2010; Weitlauf et al., 2022). A meta-analysis of differential predictors of response to ESDM in autistic children 12–48 months old found that higher pre-treatment intellectual functioning represented the strongest predictor of positive response to ESDM (Asta & Persico, 2022). A separate meta-analysis of early comprehensive treatment models for ASD (Shi et al., 2021) found that across trials examining cognitive functioning (n=420 participants), the pooled standardized mean change effect size for IQ was 0.85; only a single study (Magiati et al., 2011) demonstrated a drop in IQ whereas ten studies showed an effect size for cognitive functioning of at least 0.50. Overall, small to moderate effects on cognitive functioning were observed in autistic youth receiving early comprehensive treatment compared to treatment as usual or minimal treatment control conditions.

The significant prediction of change in autistic traits from MSEL ELC scores suggests an opportunity for early intervention to potentially modify trajectories of autistic traits from early childhood to adolescence ^1^. This relation in the ASD group may reflect that autistic children leverage cognitive skills in situations with social communication demands, whereas it may not be necessary for typically developing children to rely on cognitive skills to the same extent in such situations (i.e., social impairment in autistic individuals may be compensated by cognitive skills) (Hirosawa et al., 2020). Hus and colleagues (Hus et al., 2013) reported that multiple non-ASD-specific factors, and in particular behavior challenges, are associated with SRS-2 scores, and suggested that if these non-specific factors are not accounted for, the SRS-2 may measure general levels of impairment, including developmental difficulties and behavior problems, as well as ASD-specific symptoms, (Moody et al., 2017). In the current context, cognitive functioning in autistic and DD youth may predict the capacity to compensate for social impairments during development and/or may indicate the presence of other impairments that interfere with social development. As performance on the MSEL is related to the capacity to maintain engagement with the assessment (Akshoomoff, 2006), it is also possible that the association with the MSEL-ELC reflects protective effects of other related factors, such as attention and emotion regulation skills, on the trajectory of autistic traits. Autistic individuals with higher IQ may also engage with intervention services to a greater extent than those with lower IQ (Jonkman et al., 2023), suggesting an indirect mechanism by which higher MSEL-ELC scores in childhood are associated with an improved trajectory of autistic traits over time.

Given evidence of linkages between inflammatory processes and ASD risk (Greene et al., 2019; Hughes et al., 2023), autistic traits were hypothesized to be associated with asthma, allergy, and eczema, all of which are associated with immune system functioning; however no such relations were found. The available indicators of immune system functioning in our data may not be adequately sensitive or reflect critical time windows for which exposure would influence autistic traits. Seizures were also hypothesized to be related to autistic traits at both timepoints, and this was observed only when considering the total youth or adolescent sample. Seizures become more common with age and few families (3.8%) reported seizures at the childhood assessment, which limits our statistical power to detect associations.

Several design features should be considered when interpreting results from this study. Psychiatric symptoms and health conditions were reported by children’s caregivers and were reported when children were relatively young. This may account for the relatively lower rates of reported psychiatric symptoms relative to reports using structured clinical interviews or other diagnostic assessments and did not allow an investigation of symptom severity (Mosner et al., 2019; Stadnick et al., 2017). The age range of the sample also precluded an examination of co-occurring conditions that typically emerge later in development (e.g., psychotic disorders and substance use disorders). Likewise, the degree of functional impairment caused by parent-reported psychiatric symptoms in SEED cannot be determined. It should be noted that the measure of autistic traits, SRS-2 scores, is not a proxy for an ASD diagnosis, and a formal evaluation for ASD was not conducted at the adolescent timepoint. There was also limited information available about therapies children received between assessments. Additionally, with two waves of data collection we were unable to evaluate non-linear variability in autistic traits over time, and we could not distinguish whether conditions and symptoms associated with autistic traits share an etiologic origin with ASD (Hawks & Constantino, 2020; Rubenstein & Bishop-Fitzpatrick, 2019).

It is also noteworthy that, compared to participants without adolescent data, participants in the adolescent sample were younger at the childhood timepoint, more likely to be female, had lower SRS-2 scores, had caregivers with more education and higher income, and had a greater proportion whose mothers identified as white. It is possible that these characteristics that are related to attrition rates between the childhood and adolescent timepoints influenced the observed relations between SRS-2 scores, MSEL ELC scores, and anxiety. Finally, the models showing longitudinal relations between childhood Mullen ELC and anxiety symptoms at childhood and change in SRS-2 scores between childhood and adolescence did not control for Mullen ELC and anxiety symptoms in adolescence because these measures were not administered at the adolescent timepoint. It is thus possible that the persistence of cognitive impairments and anxiety from childhood to adolescence are influencing changes in SRS-2 scores.

Despite these limitations, this study represents the first examination of predictors of change in autistic traits in a large population-based cohort in the context of a case-control design. The substantial overlap in significant predictors of autistic traits in the ASD and POP groups underscores the potential importance of early detection and treatment of medical, developmental, and psychiatric conditions early in development potentially to improve social communication in children with and without ASD. These results contribute to our understanding of the effects of co-occurring developmental, psychiatric, behavioral, and medical symptoms and conditions on the emergence of autistic traits and underscore the need for early detection of these factors to modify trajectories social communication during development.

## Ethics Approval

### Declarations

#### Competing interests

The authors have no relevant financial or non-financial interests to disclose.

## Acknowledgements

This work was supported by Centers for Disease Control and Prevention (CDC) Cooperative Agreements announced under the following RFAs: 01086, 02199, DD11-002, DD06-003, DD04-001, and DD09-002, and by the National Institute of Environmental Health Sciences grants T32ES007018 and P30ES010126 and the National Institute of Neurological Disorders and Stroke grant K01NS099343. The findings and conclusions in this manuscript are those of the authors and do not necessarily represent the official position of the Centers for Disease Control and Prevention.

**<u>SEED 1:</u>** This study was supported by six cooperative agreements from the Centers for Disease Control and Prevention (CDC):

Cooperative Agreement Number U01DD000180 (Lisa Miller), Colorado Department of Public Health;

Cooperative Agreement Number U01DD000181 (Lisa Croen), Kaiser Foundation Research Institute;

Cooperative Agreement Number U01DD000182 (Jennifer Pinto-Martin), University of Pennsylvania;

Cooperative Agreement Number U01DD000183 (M. Daniele Fallin), Johns Hopkins University; Cooperative Agreement Number U01DD000184 (Julie Daniels), University of North Carolina at Chapel Hill; and

Cooperative Agreement Number U01DD000498 (Phil Reed), Michigan State University.

**<u>SEED Teen</u>:** This study was supported by a cooperative agreement from the Centers for Disease Control and Prevention (CDC): Cooperative Agreement Number U01DD001207 (Julie Daniels), University of North Carolina at Chapel Hill. We thank the SEED Data Coordinating Center team at the Clinical and Translational Sciences Institute of Michigan State University for their support throughout this study.

We thank all SEED participants and families for their contributions to this research. We would also like to thank the SEED Data Coordinating Center team at the Clinical and Translational Sciences Institute of Michigan State University for their technical support throughout this study. We are deeply indebted to Lisa Wiggins, Ph.D. for assistance with multiple aspect of this project.

## Authors’ contributions

KFG performed all analyses. GSD, JLD, CCB, KFG, and KKC wrote the first draft of the manuscript. All authors commented on previous versions of the manuscript. All authors read and approved the final manuscript.

## Use of identity-first language

We are using identity-first language rather than person-first language (e.g., *autistic child* rather than *child with autism*) given the current preferences of many autism stakeholders for identity-first language (Taboas et al., 2023). We recognize that language and preferences are dynamic and that not all stakeholders will share this view.

## Data availability

Data from the study described in this manuscript will be shared with National Database for Autism Research (NDAR) for those participants who gave explicit permission for data sharing. This sharing has not yet occurred.

## Footnotes

1 This possibility may not be evaluated in SEED because sufficiently detailed information on early intervention is not available.

## Notes

### Competing Interest Statement

The authors have declared no competing interest.

### Author Declarations

Institutional review boards at CDC and each site approved the SEED protocol

## References

1. Achenbach, T. M., & Rescorla, L. A. (2000). Manual for the ASEBA preschool forms and profiles (Vol. 30). Burlington, VT: University of Vermont, Research center for children, youth ….

2. Akshoomoff, N. (2006). Use of the Mullen Scales of Early Learning for the assessment of young children with Autism Spectrum Disorders. Child Neuropsychol, 12(4-5), 269–277. 10.1080/09297040500473714

3. Al-Beltagi, M. (2021). Autism medical comorbidities. World J Clin Pediatr, 10(3), 15–28. 10.5409/wjcp.v10.i3.15

4. Aldinger, K. A., Lane, C. J., Veenstra-VanderWeele, J., & Levitt, P. (2015). Patterns of Risk for Multiple Co-Occurring Medical Conditions Replicate Across Distinct Cohorts of Children with Autism Spectrum Disorder. Autism Res, 8(6), 771–781. 10.1002/aur.1492

5. Alexeeff, S. E., Yau, V., Qian, Y., Davignon, M., Lynch, F., Crawford, P., Davis, R., & Croen, L. A. (2017). Medical Conditions in the First Years of Life Associated with Future Diagnosis of ASD in Children. J Autism Dev Disord, 47(7), 2067–2079. 10.1007/s10803-017-3130-4

6. American Psychiatric, A. (2013). Diagnostic and Statistical Manual of Mental Disorders (DSM-5®).

7. Asta, L., & Persico, A. M. (2022). Differential Predictors of Response to Early Start Denver Model vs. Early Intensive Behavioral Intervention in Young Children with Autism Spectrum Disorder: A Systematic Review and Meta-Analysis. Brain Sciences, 12(11), 1499. https://www.mdpi.com/2076-3425/12/11/1499

8. August, S. M., Kiwanuka, J. N., McMahon, R. P., & Gold, J. M. (2012). The MATRICS Consensus Cognitive Battery (MCCB): clinical and cognitive correlates. Schizophr Res, 134(1), 76–82. 10.1016/j.schres.2011.10.015

9. Bishop, S. L., Guthrie, W., Coffing, M., & Lord, C. (2011). Convergent validity of the Mullen Scales of Early Learning and the differential ability scales in children with autism spectrum disorders. Am J Intellect Dev Disabil, 116(5), 331–343. 10.1352/1944-7558-116.5.331

10. Bresnahan, M., Hornig, M., Schultz, A. F., Gunnes, N., Hirtz, D., Lie, K. K., Magnus, P., Reichborn-Kjennerud, T., Roth, C., Schjølberg, S., Stoltenberg, C., Surén, P., Susser, E., & Lipkin, W. I. (2015). Association of maternal report of infant and toddler gastrointestinal symptoms with autism: evidence from a prospective birth cohort. JAMA Psychiatry, 72(5), 466–474. 10.1001/jamapsychiatry.2014.3034

11. Bryson, S. A., Corrigan, S. K., McDonald, T. P., & Holmes, C. (2008). Characteristics of children with autism spectrum disorders who received services through community mental health centers. Autism, 12(1), 65–82. 10.1177/1362361307085214

12. Canitano, R. (2007). Epilepsy in autism spectrum disorders. Eur Child Adolesc Psychiatry, 16(1), 61–66. 10.1007/s00787-006-0563-2

13. Castelbaum, L., Sylvester, C. M., Zhang, Y., Yu, Q., & Constantino, J. N. (2020). On the Nature of Monozygotic Twin Concordance and Discordance for Autistic Trait Severity: A Quantitative Analysis. Behav Genet, 50(4), 263–272. 10.1007/s10519-019-09987-2

14. Close, H. A., Lee, L. C., Kaufmann, C. N., & Zimmerman, A. W. (2012). Co-occurring conditions and change in diagnosis in autism spectrum disorders. Pediatrics, 129(2), e305–316. 10.1542/peds.2011-1717

15. Connery, K., Tippett, M., Delhey, L. M., Rose, S., Slattery, J. C., Kahler, S. G., Hahn, J., Kruger, U., Cunningham, M. W., Shimasaki, C., & Frye, R. E. (2018). Intravenous immunoglobulin for the treatment of autoimmune encephalopathy in children with autism. Transl Psychiatry, 8(1), 148. 10.1038/s41398-018-0214-7

16. Constantino, J. N. (2011). The quantitative nature of autistic social impairment. Pediatr Res, 69(5 Pt 2), 55r–62r. 10.1203/PDR.0b013e318212ec6e

17. Constantino, J. N. (2021). New guidance to seekers of autism biomarkers: an update from studies of identical twins. Mol Autism, 12(1), 28. 10.1186/s13229-021-00434-w

18. Constantino, J. N., & Gruber, C. P. (2012). Social Responsiveness Scale–Second Edition (SRS-2). In. Torrance, CA: Western Psychological Services.

19. Cummings, J. R., Lynch, F. L., Rust, K. C., Coleman, K. J., Madden, J. M., Owen-Smith, A. A., Yau, V. M., Qian, Y., Pearson, K. A., Crawford, P. M., Massolo, M. L., Quinn, V. P., & Croen, L. A. (2016). Health Services Utilization Among Children With and Without Autism Spectrum Disorders. J Autism Dev Disord, 46(3), 910–920. 10.1007/s10803-015-2634-z

20. Dawson, G., Rogers, S., Munson, J., Smith, M., Winter, J., Greenson, J., Donaldson, A., & Varley, J. (2010). Randomized, controlled trial of an intervention for toddlers with autism: the Early Start Denver Model. Pediatrics, 125(1), e17–23. 10.1542/peds.2009-0958

21. de Bruin, E. I., Ferdinand, R. F., Meester, S., de Nijs, P. F., & Verheij, F. (2007). High rates of psychiatric co-morbidity in PDD-NOS. J Autism Dev Disord, 37(5), 877–886. 10.1007/s10803-006-0215-x

22. Devitt, N. M., Gallagher, L., & Reilly, R. B. (2015). Autism Spectrum Disorder (ASD) and Fragile X Syndrome (FXS): Two Overlapping Disorders Reviewed through Electroencephalography-What Can be Interpreted from the Available Information? Brain Sci, 5(2), 92–117. 10.3390/brainsci5020092

23. Devnani, P. A., & Hegde, A. U. (2015). Autism and sleep disorders. J Pediatr Neurosci, 10(4), 304–307. 10.4103/1817-1745.174438

24. Eigsti, I.-M., Fein, D., & Larson, C. Editorial Perspective: Another look at ‘optimal outcome’ in autism spectrum disorder. Journal of Child Psychology and Psychiatry, n/a(n/a). 10.1111/jcpp.13658

25. Estes, A., Zwaigenbaum, L., Gu, H., St John, T., Paterson, S., Elison, J. T., Hazlett, H., Botteron, K., Dager, S. R., Schultz, R. T., Kostopoulos, P., Evans, A., Dawson, G., Eliason, J., Alvarez, S., Piven, J., & network, I. (2015). Behavioral, cognitive, and adaptive development in infants with autism spectrum disorder in the first 2 years of life. J Neurodev Disord, 7(1), 24. 10.1186/s11689-015-9117-6

26. Fein, D., Barton, M., Eigsti, I. M., Kelley, E., Naigles, L., Schultz, R. T., Stevens, M., Helt, M., Orinstein, A., Rosenthal, M., Troyb, E., & Tyson, K. (2013). Optimal outcome in individuals with a history of autism. J Child Psychol Psychiatry, 54(2), 195–205. 10.1111/jcpp.12037

27. Fountain, C., Winter, A. S., Cheslack-Postava, K., & Bearman, P. S. (2023). Developmental Trajectories of Autism. Pediatrics, 152(3). 10.1542/peds.2022-058674

28. Frye, R. E., Casanova, M. F., Fatemi, S. H., Folsom, T. D., Reutiman, T. J., Brown, G. L., Edelson, S. M., Slattery, J. C., & Adams, J. B. (2016). Neuropathological Mechanisms of Seizures in Autism Spectrum Disorder. Front Neurosci, 10, 192. 10.3389/fnins.2016.00192

29. Fulceri, F., Morelli, M., Santocchi, E., Cena, H., Del Bianco, T., Narzisi, A., Calderoni, S., & Muratori, F. (2016). Gastrointestinal symptoms and behavioral problems in preschoolers with Autism Spectrum Disorder. Dig Liver Dis, 48(3), 248–254. 10.1016/j.dld.2015.11.026

30. Gjevik, E., Eldevik, S., Fjaeran-Granum, T., & Sponheim, E. (2011). Kiddie-SADS reveals high rates of DSM-IV disorders in children and adolescents with autism spectrum disorders. J Autism Dev Disord, 41(6), 761–769. 10.1007/s10803-010-1095-7

31. Gotham, K., Risi, S., Pickles, A., & Lord, C. (2007). The Autism Diagnostic Observation Schedule: revised algorithms for improved diagnostic validity. J Autism Dev Disord, 37(4), 613–627. 10.1007/s10803-006-0280-1

32. Greene, R. K., Walsh, E., Mosner, M. G., & Dichter, G. S. (2019). A potential mechanistic role for neuroinflammation in reward processing impairments in autism spectrum disorder. Biol Psychol, 142, 1–12. 10.1016/j.biopsycho.2018.12.008

33. Harstad, E., Golden, M., Sideridis, G., Brewster, S. J., & Barbaresi, W. (2024). Developmental and Psychiatric Conditions Among 5-7 Year Old Children with Non-persistent and Persistent Autism. J Autism Dev Disord. 10.1007/s10803-024-06628-4

34. Harstad, E., Hanson, E., Brewster, S. J., DePillis, R., Milliken, A. L., Aberbach, G., Sideridis, G., & Barbaresi, W. J. (2023). Persistence of Autism Spectrum Disorder From Early Childhood Through School Age. JAMA Pediatrics. 10.1001/jamapediatrics.2023.4003

35. Haslam, N. (2002). Kinds of Kinds: A Conceptual Taxonomy of Psychiatric Categories. *Philosophy, Psychiatry*, & Psychology, 9(3), 203–217. 10.1353/ppp.2003.0043

36. Hawks, Z. W., & Constantino, J. N. (2020). Neuropsychiatric “Comorbidity” as Causal Influence in Autism. J Am Acad Child Adolesc Psychiatry, 59(2), 229–235. 10.1016/j.jaac.2019.07.008

37. Hirosawa, T., Kontani, K., Fukai, M., Kameya, M., Soma, D., Hino, S., Kitamura, T., Hasegawa, C., An, K. M., Takahashi, T., Yoshimura, Y., & Kikuchi, M. (2020). Different associations between intelligence and social cognition in children with and without autism spectrum disorders. PLoS One, 15(8), e0235380. 10.1371/journal.pone.0235380

38. Hughes, H. K., Moreno, R. J., & Ashwood, P. (2023). Innate immune dysfunction and neuroinflammation in autism spectrum disorder (ASD). Brain Behav Immun, 108, 245–254. 10.1016/j.bbi.2022.12.001

39. Hus, V., Bishop, S., Gotham, K., Huerta, M., & Lord, C. (2013). Factors influencing scores on the social responsiveness scale. J Child Psychol Psychiatry, 54(2), 216–224. 10.1111/j.1469-7610.2012.02589.x

40. Isaksson, J., Taylor, M. J., Lundin, K., Neufeld, J., & Bölte, S. (2019). Familial confounding on the ability to read minds: A co-twin control study. Autism, 23(8), 1948–1956. 10.1177/1362361319836380

41. Jonkman, K. M., Back, E., & Begeer, S. (2023). Predicting intervention use in autistic children: Demographic and autism-specific characteristics. Autism, 27(2), 428–442. 10.1177/13623613221102748

42. Joshi, G., Petty, C., Wozniak, J., Henin, A., Fried, R., Galdo, M., Kotarski, M., Walls, S., & Biederman, J. (2010). The heavy burden of psychiatric comorbidity in youth with autism spectrum disorders: a large comparative study of a psychiatrically referred population. J Autism Dev Disord, 40(11), 1361–1370. 10.1007/s10803-010-0996-9

43. Jyonouchi, H. (2010). Autism spectrum disorders and allergy: observation from a pediatric allergy/immunology clinic. Expert Rev Clin Immunol, 6(3), 397–411. 10.1586/eci.10.18

44. Kamio, Y., Inada, N., & Koyama, T. (2013). A nationwide survey on quality of life and associated factors of adults with high-functioning autism spectrum disorders. Autism, 17(1), 15–26. 10.1177/1362361312436848

45. Kanne, S. M., Abbacchi, A. M., & Constantino, J. N. (2009). Multi-informant ratings of psychiatric symptom severity in children with autism spectrum disorders: the importance of environmental context. J Autism Dev Disord, 39(6), 856–864. 10.1007/s10803-009-0694-7

46. Khachadourian, V., Mahjani, B., Sandin, S., Kolevzon, A., Buxbaum, J. D., Reichenberg, A., & Janecka, M. (2023). Comorbidities in autism spectrum disorder and their etiologies. Transl Psychiatry, 13(1), 71. 10.1038/s41398-023-02374-w

47. Kim, S. H., & Byun, Y. (2018). Trajectories of Symptom Clusters, Performance Status, and Quality of Life During Concurrent Chemoradiotherapy in Patients With High-Grade Brain Cancers. Cancer Nurs, 41(1), E38–E47. 10.1097/NCC.0000000000000435

48. Klaiman, C., White, S., Richardson, S., McQueen, E., Walum, H., Aoki, C., Smith, C., Minjarez, M., Bernier, R., Pedapati, E., Bishop, S., Ence, W., Wainer, A., Moriuchi, J., Tay, S.-W., Deng, Y., Jones, W., Gillespie, S., & Klin, A. (2022). Expert Clinician Certainty in Diagnosing Autism Spectrum Disorder in 16–30-Month-Olds: A Multi-site Trial Secondary Analysis. Journal of Autism and Developmental Disorders. 10.1007/s10803-022-05812-8

49. Klukowski, M., Wasilewska, J., & Lebensztejn, D. (2015). Sleep and gastrointestinal disturbances in autism spectrum disorder in children. Dev Period Med, 19(2), 157–161.

50. Kraper, C. K., Kenworthy, L., Popal, H., Martin, A., & Wallace, G. L. (2017). The Gap Between Adaptive Behavior and Intelligence in Autism Persists into Young Adulthood and is Linked to Psychiatric Co-morbidities. J Autism Dev Disord, 47(10), 3007–3017. 10.1007/s10803-017-3213-2

51. Lai, M. C., Kassee, C., Besney, R., Bonato, S., Hull, L., Mandy, W., Szatmari, P., & Ameis, S. H. (2019). Prevalence of co-occurring mental health diagnoses in the autism population: a systematic review and meta-analysis. Lancet Psychiatry, 6(10), 819–829. 10.1016/S2215-0366(19)30289-5

52. Lamb, G. V., Green, R. J., & Olorunju, S. (2019). Tracking epilepsy and autism. *The Egyptian Journal of Neurology*, Psychiatry and Neurosurgery, 55(1), 55. 10.1186/s41983-019-0103-x

53. Landa, R. J., Reetzke, R., Holingue, C. B., Herman, D., & Hess, C. R. (2022). Diagnostic Stability and Phenotypic Differences Among School-Age Children Diagnosed With ASD Before Age 2. Front Psychiatry, 13, 805686. 10.3389/fpsyt.2022.805686

54. Leyfer, O. T., Folstein, S. E., Bacalman, S., Davis, N. O., Dinh, E., Morgan, J., Tager-Flusberg, H., & Lainhart, J. E. (2006). Comorbid psychiatric disorders in children with autism: interview development and rates of disorders. J Autism Dev Disord, 36(7), 849–861. 10.1007/s10803-006-0123-0

55. Lord, C. (2018). For Better or for Worse? Later Diagnoses of Autism Spectrum Disorder in Some Younger Siblings of Already Diagnosed Children. J Am Acad Child Adolesc Psychiatry, 57(11), 822–823. 10.1016/j.jaac.2018.08.008

56. Lord, C., Rutter, M., DiLavore, P., & Risi, S. (1999). Autism diagnostic observation schedule. In. Los Angeles: Western Psychological Services.

57. Lord, C., Rutter, M., & Le Couteur, A. (1994). Autism Diagnostic Interview-Revised: a revised version of a diagnostic interview for caregivers of individuals with possible pervasive developmental disorders. J Autism Dev Disord, 24(5), 659–685. 10.1007/BF02172145

58. Maenner, M. J., Warren, Z., Williams, A. R., & al., e. (2023). Prevalence and Characteristics of Autism Spectrum Disorder Among Children Aged 8 Years — Autism and Developmental Disabilities Monitoring Network, 11 Sites, United States, 2020. MMWR Surveill Summ, 2023;72(No. SS-2), 1–14.

59. Magiati, I., Moss, J., Charman, T., & Howlin, P. (2011). Patterns of change in children with autism spectrum disorders who received community based comprehensive interventions in their pre-school years: A seven year follow-up study. Research in Autism Spectrum Disorders, 5(3), 1016–1027. 10.1016/j.rasd.2010.11.007

60. Malow, B. A., Qian, Y., Ames, J. L., Alexeeff, S., & Croen, L. A. (2023). Health conditions in autism: Defining the trajectory from adolescence to early adulthood. Autism Research, 16(7), 1437–1449. 10.1002/aur.2960

61. McDougle, C. J., Stigler, K. A., & Posey, D. J. (2003). Treatment of aggression in children and adolescents with autism and conduct disorder. J Clin Psychiatry, 64 *Suppl 4*, 16–25. https://www.ncbi.nlm.nih.gov/pubmed/12672261

62. http://www.psychiatrist.com/jcp/article/pages/2003/v64s04/v64s0403.aspx

63. Mohn, C., Sundet, K., & Rund, B. R. (2014). The relationship between IQ and performance on the MATRICS consensus cognitive battery. Schizophr Res Cogn, 1(2), 96–100. 10.1016/j.scog.2014.06.003

64. Moody, E. J., Reyes, N., Ledbetter, C., Wiggins, L., DiGuiseppi, C., Alexander, A., Jackson, S., Lee, L. C., Levy, S. E., & Rosenberg, S. A. (2017). Screening for Autism with the SRS and SCQ: Variations across Demographic, Developmental and Behavioral Factors in Preschool Children. J Autism Dev Disord, 47(11), 3550–3561. 10.1007/s10803-017-3255-5

65. Morrison, K. E., Pinkham, A. E., Kelsven, S., Ludwig, K., Penn, D. L., & Sasson, N. J. (2019). Psychometric Evaluation of Social Cognitive Measures for Adults with Autism. Autism Res, 12(5), 766–778. 10.1002/aur.2084

66. Mosner, M. G., Kinard, J. L., Shah, J. S., McWeeny, S., Greene, R. K., Lowery, S. C., Mazefsky, C. A., & Dichter, G. S. (2019). Rates of Co-occurring Psychiatric Disorders in Autism Spectrum Disorder Using the Mini International Neuropsychiatric Interview. J Autism Dev Disord, 49(9), 3819–3832. 10.1007/s10803-019-04090-1

67. Moulton, E., Barton, M., Robins, D. L., Abrams, D. N., & Fein, D. (2016). Early Characteristics of Children with ASD Who Demonstrate Optimal Progress Between Age Two and Four. J Autism Dev Disord, 46(6), 2160–2173. 10.1007/s10803-016-2745-1

68. Mullen, E. M. (1995). Mullen scales of early learning (AGS ed.). In. Circle Pines, MN: American Guidance Service Inc.

69. Murray, M., & Blume, J. (2021). FDRestimation: Flexible False Discovery Rate Computation in R [version 2; peer review: 2 approved]. F1000Research, 10(441). 10.12688/f1000research.52999.2

70. Owens, J. A., Spirito, A., & McGuinn, M. (2000). The Children’s Sleep Habits Questionnaire (CSHQ): psychometric properties of a survey instrument for school-aged children. Sleep, 23(8), 1043–1051. https://www.ncbi.nlm.nih.gov/pubmed/11145319

71. Oxelgren, U. W., Myrelid, Å., Annerén, G., Ekstam, B., Göransson, C., Holmbom, A., Isaksson, A., Åberg, M., Gustafsson, J., & Fernell, E. (2017). Prevalence of autism and attention-deficit-hyperactivity disorder in Down syndrome: a population-based study. Dev Med Child Neurol, 59(3), 276–283. 10.1111/dmcn.13217

72. Ozonoff, S., Young, G. S., Belding, A., Hill, M., Hill, A., Hutman, T., Johnson, S., Miller, M., Rogers, S. J., Schwichtenberg, A. J., Steinfeld, M., & Iosif, A. M. (2014). The broader autism phenotype in infancy: when does it emerge? J Am Acad Child Adolesc Psychiatry, 53(4), 398–407 e392. 10.1016/j.jaac.2013.12.020

73. Ozonoff, S., Young, G. S., Brian, J., Charman, T., Shephard, E., Solish, A., & Zwaigenbaum, L. (2018). Diagnosis of Autism Spectrum Disorder After Age 5 in Children Evaluated Longitudinally Since Infancy. J Am Acad Child Adolesc Psychiatry, 57(11), 849–857 e842. 10.1016/j.jaac.2018.06.022

74. Ozonoff, S., Young, G. S., Landa, R. J., Brian, J., Bryson, S., Charman, T., Chawarska, K., Macari, S. L., Messinger, D., Stone, W. L., Zwaigenbaum, L., & Iosif, A. M. (2015). Diagnostic stability in young children at risk for autism spectrum disorder: a baby siblings research consortium study. J Child Psychol Psychiatry, 56(9), 988–998. 10.1111/jcpp.12421

75. Pacheva, I., Ivanov, I., Yordanova, R., Gaberova, K., Galabova, F., Panova, M., Petkova, A., Timova, E., & Sotkova, I. (2019). Epilepsy in Children with Autistic Spectrum Disorder. Children (Basel), 6(2). 10.3390/children6020015

76. Peterson, C. C., Slaughter, V., & Brownell, C. (2015). Children with autism spectrum disorder are skilled at reading emotion body language. J Exp Child Psychol, 139, 35–50. 10.1016/j.jecp.2015.04.012

77. Pierce, K., Gazestani, V. H., Bacon, E., Barnes, C. C., Cha, D., Nalabolu, S., Lopez, L., Moore, A., Pence-Stophaeros, S., & Courchesne, E. (2019). Evaluation of the Diagnostic Stability of the Early Autism Spectrum Disorder Phenotype in the General Population Starting at 12 Months. JAMA Pediatr, 173(6), 578–587. 10.1001/jamapediatrics.2019.0624

78. Powell, P. S., Pazol, K., Wiggins, L. D., Daniels, J. L., Dichter, G. S., Bradley, C. B., Pretzel, R., Kloetzer, J., McKenzie, C., Scott, A., Robinson, B., Sims, A. S., Kasten, E. P., Fallin, M. D., Levy, S. E., Dietz, P. M., & Cogswell, M. E. (2021). Health Status and Health Care Use Among Adolescents Identified With and Without Autism in Early Childhood - Four U.S. Sites, 2018-2020. MMWR Morb Mortal Wkly Rep, 70(17), 605–611. 10.15585/mmwr.mm7017a1

79. Prosperi, M., Santocchi, E., Muratori, F., Narducci, C., Calderoni, S., Tancredi, R., Morales, M. A., & Guiducci, L. (2019). Vocal and motor behaviors as a possible expression of gastrointestinal problems in preschoolers with Autism Spectrum Disorder. BMC Pediatr, 19(1), 466. 10.1186/s12887-019-1841-8

80. Reynolds, A. M., Soke, G. N., Sabourin, K. R., Hepburn, S., Katz, T., Wiggins, L. D., Schieve, L. A., & Levy, S. E. (2019). Sleep Problems in 2- to 5-Year-Olds With Autism Spectrum Disorder and Other Developmental Delays. Pediatrics, 143(3). 10.1542/peds.2018-0492

81. Rosen, T. E., Mazefsky, C. A., Vasa, R. A., & Lerner, M. D. (2018). Co-occurring psychiatric conditions in autism spectrum disorder. Int Rev Psychiatry, 30(1), 40–61. 10.1080/09540261.2018.1450229

82. Rubenstein, E., & Bishop-Fitzpatrick, L. (2019). A matter of time: The necessity of temporal language in research on health conditions that present with autism spectrum disorder. Autism Res, 12(1), 20–25. 10.1002/aur.2010

83. Rutter, M., Bailey, A., & Lord, C. (2003). SCQ: Social communication questionnaire. In. Los Angeles: Western Psychological Services.

84. Sasson, N. J., Morrison, K. E., Kelsven, S., & Pinkham, A. E. (2020). Social cognition as a predictor of functional and social skills in autistic adults without intellectual disability. Autism Res, 13(2), 259–270. 10.1002/aur.2195

85. Schendel, D. E., Diguiseppi, C., Croen, L. A., Fallin, M. D., Reed, P. L., Schieve, L. A., Wiggins, L. D., Daniels, J., Grether, J., Levy, S. E., Miller, L., Newschaffer, C., Pinto-Martin, J., Robinson, C., Windham, G. C., Alexander, A., Aylsworth, A. S., Bernal, P., Bonner, J. D., . . . Yeargin-Allsopp, M. (2012). The Study to Explore Early Development (SEED): a multisite epidemiologic study of autism by the Centers for Autism and Developmental Disabilities Research and Epidemiology (CADDRE) network. J Autism Dev Disord, 42(10), 2121–2140. 10.1007/s10803-012-1461-8

86. Shi, B., Wu, W., Dai, M., Zeng, J., Luo, J., Cai, L., Wan, B., & Jing, J. (2021). Cognitive, Language, and Behavioral Outcomes in Children With Autism Spectrum Disorders Exposed to Early Comprehensive Treatment Models: A Meta-Analysis and Meta-Regression. Front Psychiatry, 12, 691148. 10.3389/fpsyt.2021.691148

87. Shultz, S., Klin, A., & Jones, W. (2018). Neonatal Transitions in Social Behavior and Their Implications for Autism. Trends Cogn Sci, 22(5), 452–469. 10.1016/j.tics.2018.02.012

88. Simonoff, E., Pickles, A., Charman, T., Chandler, S., Loucas, T., & Baird, G. (2008). Psychiatric disorders in children with autism spectrum disorders: prevalence, comorbidity, and associated factors in a population-derived sample. J Am Acad Child Adolesc Psychiatry, 47(8), 921–929. 10.1097/CHI.0b013e318179964f

89. Soke, G. N., Maenner, M. J., Christensen, D., Kurzius-Spencer, M., & Schieve, L. A. (2018). Prevalence of Co-occurring Medical and Behavioral Conditions/Symptoms Among 4- and 8-Year-Old Children with Autism Spectrum Disorder in Selected Areas of the United States in 2010. J Autism Dev Disord, 48(8), 2663–2676. 10.1007/s10803-018-3521-1

90. Stadnick, N., Chlebowski, C., Baker-Ericzén, M., Dyson, M., Garland, A., & Brookman-Frazee, L. (2017). Psychiatric comorbidity in autism spectrum disorder: Correspondence between mental health clinician report and structured parent interview. Autism, 21(7), 841–851. 10.1177/1362361316654083

91. Su, T. P., Chen, M. H., & Tu, P. C. (2022). Using big data of genetics, health claims, and brain imaging to challenge the categorical classification in mental illness. J Chin Med Assoc, 85(2), 139–144. 10.1097/jcma.0000000000000675

92. Taboas, A., Doepke, K., & Zimmerman, C. (2023). Preferences for identity-first versus person-first language in a US sample of autism stakeholders. Autism, 27(2), 565–570. 10.1177/13623613221130845

93. Team, R. C. (2013). A Language and Environment for Statistical Computing. In. Vienna: A Language and Environment for Statistical Computing.

94. Tunç, B., Pandey, J., St John, T., Meera, S. S., Maldarelli, J. E., Zwaigenbaum, L., Hazlett, H. C., Dager, S. R., Botteron, K. N., Girault, J. B., McKinstry, R. C., Verma, R., Elison, J. T., Pruett, J. R., Jr., Piven, J., Estes, A. M., & Schultz, R. T. (2021). Diagnostic shifts in autism spectrum disorder can be linked to the fuzzy nature of the diagnostic boundary: a data-driven approach. J Child Psychol Psychiatry, 62(10), 1236–1245. 10.1111/jcpp.13406

95. Tunc, B., Pandey, J., St John, T., Meera, S. S., Maldarelli, J. E., Zwaigenbaum, L., Hazlett, H. C., Dager, S. R., Botteron, K. N., Girault, J. B., McKinstry, R. C., Verma, R., Elison, J. T., Pruett, J. R., Jr., Piven, J., Estes, A. M., Schultz, R. T., & Network, I. (2021). Diagnostic shifts in autism spectrum disorder can be linked to the fuzzy nature of the diagnostic boundary: a data-driven approach. J Child Psychol Psychiatry. 10.1111/jcpp.13406

96. Wagner, R. E., Zhang, Y., Gray, T., Abbacchi, A., Cormier, D., Todorov, A., & Constantino, J. N. (2019). Autism-Related Variation in Reciprocal Social Behavior: A Longitudinal Study. Child Dev, 90(2), 441–451. 10.1111/cdev.13170

97. Wasilewska, J., & Klukowski, M. (2015). Gastrointestinal symptoms and autism spectrum disorder: links and risks - a possible new overlap syndrome. Pediatric Health Med Ther, 6, 153–166. 10.2147/phmt.S85717

98. Weber, R. J., & Gadow, K. D. (2017). Relation of Psychiatric Symptoms with Epilepsy, Asthma, and Allergy in Youth with ASD vs. Psychiatry Referrals. J Abnorm Child Psychol, 45(6), 1247–1257. 10.1007/s10802-016-0212-2

99. Weitlauf, A. S., Broderick, N., Alacia Stainbrook, J., Slaughter, J. C., Taylor, J. L., Herrington, C. G., Nicholson, A. G., Santulli, M., Dorris, K., Garrett, L. J., Hopton, M., Kinsman, A., Morton, M., Vogel, A., Dykens, E. M., Pablo Juárez, A., & Warren, Z. E. (2022). A Longitudinal RCT of P-ESDM With and Without Parental Mindfulness Based Stress Reduction: Impact on Child Outcomes. J Autism Dev Disord, 52(12), 5403–5413. 10.1007/s10803-021-05399-6

100. Wiggins, L. D., Levy, S. E., Daniels, J., Schieve, L., Croen, L. A., DiGuiseppi, C., Blaskey, L., Giarelli, E., Lee, L. C., Pinto-Martin, J., Reynolds, A., Rice, C., Rosenberg, C. R., Thompson, P., Yeargin-Allsopp, M., Young, L., & Schendel, D. (2015). Autism spectrum disorder symptoms among children enrolled in the Study to Explore Early Development (SEED). J Autism Dev Disord, 45(10), 3183–3194. 10.1007/s10803-015-2476-8

101. Wiggins, L. D., Reynolds, A., Rice, C. E., Moody, E. J., Bernal, P., Blaskey, L., Rosenberg, S. A., Lee, L. C., & Levy, S. E. (2015). Using standardized diagnostic instruments to classify children with autism in the study to explore early development. J Autism Dev Disord, 45(5), 1271–1280. 10.1007/s10803-014-2287-3

102. Yang, X. L., Liang, S., Zou, M. Y., Sun, C. H., Han, P. P., Jiang, X. T., Xia, W., & Wu, L. J. (2018). Are gastrointestinal and sleep problems associated with behavioral symptoms of autism spectrum disorder? Psychiatry Res, 259, 229–235. 10.1016/j.psychres.2017.10.040

103. Zimmerman, A. W., Connors, S. L., Matteson, K. J., Lee, L. C., Singer, H. S., Castaneda, J. A., & Pearce, D. A. (2007). Maternal antibrain antibodies in autism. Brain Behav Immun, 21(3), 351–357. 10.1016/j.bbi.2006.08.005

104. Zwaigenbaum, L., Bryson, S. E., Brian, J., Smith, I. M., Roberts, W., Szatmari, P., Roncadin, C., Garon, N., & Vaillancourt, T. (2016). Stability of diagnostic assessment for autism spectrum disorder between 18 and 36 months in a high-risk cohort. Autism Res, 9(7), 790–800. 10.1002/aur.1585

