## Supplemental Information for "Changes in Autism Traits from Early Childhood to Adolescence in the Study to Explore Early Development"

Supplementary Information

Supplementary Table 1. Distributions of developmental, behavioral, and psychiatric variables and medical conditions among children (ages 2-5 years) by study group in the Study to Explore Early Development - Phase 1 (2007-2011)

| Developmental, Behavioral, and Psychiatric Predictors | | | | | | | |
| --- | --- | --- | --- | --- | --- | --- | --- |
|  | N (%) / Mean (Std Dev) | | |  | Significance Testing (χ2/t-test p-values) | | |
| Childhood Predictors | POP | ASD | DD |  | POP vs ASD | ASD vs DD | POP vs DD |
| ADHD |  |  |  |  | <0.001 | 0.723 | <0.001 |
| Yes | 7 (0.8) | 56 (8.2) | 74 (7.6) |  |  |  |  |
| No | 867 (99.2) | 625 (91.8) | 897 (92.4) |  |  |  |  |
| CBCL Externalizing t-score (mean, SD) | 43.86 (10.17) | 60.27 (11.39) | 49.87 (12.40) |  | <0.001 | <0.001 | <0.001 |
| CBCL Internalizing t-score (mean, SD) | 45.03 (10.47) | 62.52 (9.59) | 50.96 (11.98) |  | <0.001 | <0.001 | <0.001 |
| Learning Disability |  |  |  |  | <0.001 | <0.001 | <0.001 |
| Yes | 8 (0.9) | 111 (16.3) | 99 (10.2) |  |  |  |  |
| No | 866 (99.1) | 570 (83.7) | 872 (89.8) |  |  |  |  |
| Motor/Movement Problems |  |  |  |  | <0.001 | 0.009 | <0.001 |
| Yes | 13 (1.5) | 136 (20.0) | 145 (14.9) |  |  |  |  |
| No | 861 (98.5) | 545 (80.0) | 826 (85.1) |  |  |  |  |
| Mullen Early Learning Composite (mean, SD) | 102.32 (14.62) | 66.91 (20.02) | 86.33 (21.02) |  | <0.001 | <0.001 | <0.001 |
| Self-injurious behaviors |  |  |  |  | <0.001 | <0.001 | <0.001 |
| Yes | 1 (0.1) | 39 (5.7) | 20 (2.1) |  |  |  |  |
| No | 873 (99.9) | 642 (94.3) | 951 (97.9) |  |  |  |  |
| Sensory Integration Disorder |  |  |  |  | <0.001 | <0.001 | <0.001 |
| Yes | 8 (0.9) | 195 (28.6) | 104 (10.7) |  |  |  |  |
| No | 866 (99.1) | 486 (71.4) | 867 (89.3) |  |  |  |  |
| Speech Delay |  |  |  |  | <0.001 | 0.008 | <0.001 |
| Yes | 80 (9.2) | 458 (67.3) | 590 (60.8) |  |  |  |  |
| No | 794 (90.8) | 223 (32.7) | 381 (39.2) |  |  |  |  |

| Other Predictors | | | | | | | |
| --- | --- | --- | --- | --- | --- | --- | --- |
| Allergy/Eczema |  |  |  |  | <0.001 | 0.039 | 0.032 |
| Yes | 303 (35.2) | 306 (45.4) | 381 (40.1) |  |  |  |  |
| No | 559 (64.8) | 368 (54.6) | 568 (59.9) |  |  |  |  |
| Asthma |  |  |  |  | 0.167 | 0.471 | 0.018 |
| Yes | 92 (10.7) | 88 (13.1) | 136 (14.5) |  |  |  |  |
| No | 770 (89.3) | 584 (86.9) | 803 (85.5) |  |  |  |  |
| Birth Defects |  |  |  |  | 0.007 | 0.085 | <0.001 |
| Yes | 23 (2.6) | 37 (5.4) | 75 (7.7) |  |  |  |  |
| No | 851 (97.4) | 644 (94.6) | 896 (92.3) |  |  |  |  |
| GIS |  |  |  |  | <0.001 | <0.001 | <0.001 |
| Yes | 96 (11.3) | 219 (33.3) | 182 (19.5) |  |  |  |  |
| No | 757 (88.7) | 438 (66.7) | 753 (80.5) |  |  |  |  |
| Hearing Problems |  |  |  |  | <0.001 | 0.037 | <0.001 |
| Yes | 13 (1.5) | 43 (6.3) | 90 (9.3) |  |  |  |  |
| No | 861 (98.5) | 638 (93.7) | 881 (90.7) |  |  |  |  |
| Pneumonia |  |  |  |  | 0.428 | 0.041 | 0.002 |
| Yes | 42 (4.8) | 40 (5.8) | 84 (8.6) |  |  |  |  |
| No | 833 (95.2) | 646 (94.2) | 889 (91.4) |  |  |  |  |
| Respiratory Syncytial Virus |  |  |  |  | 0.975 | 0.522 | 0.576 |
| Yes | 64 (7.3) | 49 (7.1) | 79 (8.1) |  |  |  |  |
| No | 811 (92.7) | 637 (92.9) | 894 (91.9) |  |  |  |  |
| Seizure |  |  |  |  | <0.001 | 0.431 | <0.001 |
| Yes | 1 (0.1) | 26 (3.8) | 29 (3.0) |  |  |  |  |
| No | 873 (99.9) | 655 (96.2) | 942 (97.0) |  |  |  |  |
| Sleep Score (mean, Std Dev) | 45.53 (8.28) | 51.21 (11.22) | 48.54 (10.32) |  | <0.001 | <0.001 | <0.001 |

Note: POP=Population controls; ASD=autism spectrum disorder; DD=Developmental Delay; ADHD=attention deficit hyperactivity disorder; Note: CBCL= Child Behavior Checklist; GIS=gastrointestinal symptoms.

Supplementary Table 2. Distributions of psychiatric and medical conditions among adolescents (ages 12-16 years) by study group in the Study to Explore Early Development Teen (2017-2021).

| Developmental, Behavioral, and Psychiatric Conditions | | | | | | | | | | | | | | |
| --- | --- | --- | --- | --- | --- | --- | --- | --- | --- | --- | --- | --- | --- | --- |
|  | N (%) / Mean (SD) | | | | |  | Significance Testing (χ2/t-test p-values) | | | | | | | |
| Adolescent Predictors | POP | | ASD | DD | |  | POP vs ASD | | | ASD vs DD | | POP vs DD | | |
| ADHD |  | |  |  | |  | <0.001 | | | 0.069 | | <0.001 | | |
| Yes | 51 (16.3) | | 81 (38.9) | 106 (31.0) | |  |  | | |  | |  | | |
| No | 262 (83.7) | | 127 (61.1) | 236 (69.0) | |  |  | | |  | |  | | |
| Anxiety |  | |  |  | |  | <0.001 | | | 0.014 | | 0.001 | | |
| Yes | 51 (16.3) | | 78 (37.7) | 94 (27.3) | |  |  | | |  | |  | | |
| No | 262 (83.7) | | 129 (62.3) | 250 (72.7) | |  |  | | |  | |  | | |
| Behavioral Concerns |  | |  |  | |  | <0.001 | | | <0.001 | | <0.001 | | |
| Yes | 7 (2.2) | | 56 (27.2) | 31 (9.1) | |  |  | | |  | |  | | |
| No | 305 (97.8) | | 150 (72.8) | 311 (90.9) | |  |  | | |  | |  | | |
| Depression |  | |  |  | |  | 0.964 | | | 0.726 | | 0.494 | | |
| Yes | 21 (6.7) | | 15 (7.2) | 29 (8.4) | |  |  | | |  | |  | | |
| No | 292 (93.3) | | 193 (92.8) | 315 (91.6) | |  |  | | |  | |  | | |
| Developmental Concerns |  | |  |  | |  | <0.001 | | | <0.001 | | <0.001 | | |
| Yes | 6 (1.9) | | 114 (57.0) | 91 (26.9) | |  |  | | |  | |  | | |
| No | 304 (98.1) | | 86 (43.0) | 247 (73.1) | |  |  | | |  | |  | | |
| Externalizing conditions |  | |  |  | |  | <0.001 | | | <0.001 | | <0.001 | | |
| Yes | 53 (16.9) | | 107 (51.4) | 115 (33.6) | |  |  | | |  | |  | | |
| No | 260 (83.1) | | 101 (48.6) | 227 (66.4) | |  |  | | |  | |  | | |
| Internalizing conditions |  | |  |  | |  | <0.001 | | | 0.023 | | 0.002 | | |
| Yes | 58 (18.5) | | 81 (39.1) | 101 (29.4) | |  |  | | |  | |  | | |
| No | 255 (81.5) | | 126 (60.9) | 243 (70.6) | |  |  | | |  | |  | | |
| Sensory Integration Disorder |  | |  |  | |  | <0.001 | | | <0.001 | | <0.001 | | |
| Yes | 1 (0.3) | | 57 (28.5) | 29 (8.8) | |  |  | | |  | |  | | |
| No | 311 (99.7) | | 143 (71.5) | 301 (91.2) | |  |  | | |  | |  | | |
| Speech Delay |  | |  |  | |  | <0.001 | | | <0.001 | | <0.001 | | |
| Yes | 7 (2.2) | | 89 (44.1) | 79 (23.2) | |  |  | | |  | |  | | |
| No | 305 (97.8) | | 113 (55.9) | 261 (76.8) | |  |  | | |  | |  | | |
| Other Conditions | | | | | | | | | | | | | | |
| Allergy/Eczema | |  | | |  | | |  |  | | 0.027 | | 0.091 | 0.58 |
| Yes | | 140 (45.0) | | | 114 (55.3) | | | 161 (47.5) |  | |  | |  |  |
| No | | 171 (55.0) | | | 92 (44.7) | | | 178 (52.5) |  | |  | |  |  |
| Asthma | |  | | |  | | |  |  | | 0.621 | | 0.324 | 0.061 |
| Yes | | 31 (10.0) | | | 24 (11.8) | | | 52 (15.5) |  | |  | |  |  |
| No | | 278 (90.0) | | | 179 (88.2) | | | 289 (84.8) |  | |  | |  |  |
| GIS | |  | | |  | | |  |  | | 0.041 | | 0.957 | 0.012 |
| Yes | | 36 (11.5) | | | 38 (18.3) | | | 65 (18.8) |  | |  | |  |  |
| No | | 277 (88.5) | | | 170 (81.7) | | | 280 (81.2) |  | |  | |  |  |
| Parasomnia Score (mean, SD) | | 7.75 (1.12) | | | 8.40 (1.68) | | | 8.17 (1.53) |  | | <0.001 | | 0.099 | <0.001 |
| Seizure | |  | | |  | | |  |  | | <0.001 | | 0.018 | 0.061 |
| Yes | | 1 (0.3) | | | 14 (6.8) | | | 8 (2.3) |  | |  | |  |  |
| No | | 311 (99.7) | | | 191 (93.2) | | | 335 (97.7) |  | |  | |  |  |
| Self-injurious behaviors | |  | | |  | | |  |  | | <0.001 | | <0.001 | 0.482 |
| Yes | | 4 (1.3) | | | 25 (12.1) | | | 8 (2.3) |  | |  | |  |  |
| No | | 307 (98.7) | | | 182 (87.9) | | | 335 (97.7) |  | |  | |  |  |
| Sleep Duration Score (mean, SD) | | 4.18 (1.59) | | | 4.42 (1.85) | | | 4.01 (1.59) |  | | 0.11 | | 0.006 | 0.187 |

Note: POP=Population controls; ASD=autism spectrum disorder; DD=Developmental Delay; ADHD=attention deficit hyperactivity disorder; GIS=gastrointestinal symptoms.

Supplemental Table 3. Childhood (ages 2-5 years) Study to Explore Early Development – Phase 1 (2007-2011) cross-sectional analyses: associations between childhood predictors and childhood SRS-2 scores.

| Developmental, Behavioral, and Psychiatric Conditions | | | | | | | | | | | |
| --- | --- | --- | --- | --- | --- | --- | --- | --- | --- | --- | --- |
|  | POP | |  | ASD | |  | DD | |  | ~~Group Interaction~~ | |
| Predictors | Estimate | p-value |  | Estimate | p-value |  | Estimate | p-value |  | ~~F~~ | ~~p-value~~ |
| ADHD: Yes | 8.33 | 0.011 |  | 3.90 | 0.034 |  | 8.38 | <0.001 |  | ~~5.69~~ | ~~0.009~~ |
| CBCL Externalizing T-score | 4.82 | <0.001 |  | 6.37 | <0.001 |  | 7.05 | <0.001 |  | ~~11.60~~ | ~~<0.0001~~ |
| CBCL Internalizing T-score | 4.85 | <0.001 |  | 9.02 | <0.001 |  | 7.70 | <0.001 |  | ~~30.78~~ | ~~<0.0001~~ |
| Learning Disability: Yes | 6.65 | 0.011 |  | 0.17 | 0.933 |  | 8.00 | <0.001 |  | ~~13.56~~ | ~~<0.0001~~ |
| Movement or Motor Problems: Yes | 5.31 | 0.017 |  | 1.81 | 0.187 |  | 6.10 | <0.001 |  | ~~2.73~~ | ~~0.110~~ |
| Mullen ELC Standard Score | -2.70 | <0.001 |  | -2.65 | <0.001 |  | -4.28 | <0.001 |  | ~~8.82~~ | ~~0.0004~~ |
| Self-Injurious Behavior: Yes | - | - |  | 5.32 | 0.016 |  | 7.69 | 0.004 |  | ~~1.14~~ | ~~0.369~~ |
| Speech or Language Disorder: Yes | 4.00 | <0.001 |  | 0.34 | 0.809 |  | 2.19 | 0.010 |  | ~~2.35~~ | ~~0.144~~ |
| Other Conditions | | | | | | | | | | | |
| Allergy/Eczema | 1.19 | 0.035 |  | 1.63 | 0.117 |  | 0.22 | 0.818 |  | ~~0.74~~ | ~~0.540~~ |
| Asthma | 0.81 | 0.384 |  | 2.46 | 0.120 |  | 2.15 | 0.067 |  | ~~1.03~~ | ~~0.433~~ |
| Birth Defects: Yes | 0.06 | 0.987 |  | -1.81 | 0.448 |  | 1.46 | 0.354 |  | ~~0.78~~ | ~~0.536~~ |
| GIS: Yes | 2.57 | 0.002 |  | 4.39 | <0.001 |  | 3.72 | <0.001 |  | ~~1.54~~ | ~~0.287~~ |
| Hearing Problems: Yes | 5.03 | 0.024 |  | 2.79 | 0.187 |  | -1.15 | 0.424 |  | ~~3.07~~ | ~~0.082~~ |
| Pneumonia: Yes | 0.02 | 0.987 |  | 2.96 | 0.187 |  | 1.91 | 0.205 |  | ~~0.74~~ | ~~0.540~~ |
| Respiratory Syncytial Virus: Yes | -0.85 | 0.424 |  | 3.30 | 0.095 |  | 0.43 | 0.809 |  | ~~2.48~~ | ~~0.132~~ |
| Epilepsy/Seizures | 3.59 | 0.669 |  | 3.57 | 0.187 |  | 5.46 | 0.021 |  | ~~0.16~~ | ~~0.889~~ |
| Sensory Integration Disorder: Yes | 10.52 | <0.001 |  | 5.62 | <0.001 |  | 8.47 | <0.001 |  | ~~2.38~~ | ~~0.143~~ |
| CSHQ Total Sleep Score | 2.26 | <0.001 |  | 2.72 | <0.001 |  | 3.47 | <0.001 |  | ~~4.59~~ | ~~0.022~~ |

Models included group, study site, child sex, child age at the Mullen assessment, maternal age, maternal education, and maternal percent poverty level as covariates (continuous covariates are centered and scaled across diagnostic groups). Analysis of variance models were used to test for the interaction between group and each predictor while controlling for covariates. SRS=Social Responsiveness Scale; CBCL= Child Behavior Checklist; POP=population controls; ASD=autism spectrum disorder; DD=developmental delay; ADHD=attention deficit hyperactivity disorder; GIS=gastrointestinal symptoms. Effective sample size varied minimally per model because incomplete cases were dropped on a per-model basis, but the effective sample sizes did not differ by more than 5% within any model group. Dashes indicate that the sample size was too small to allow for model fitting. Estimates are regression coefficients.

Supplemental Table 4. Adolescent (ages 12-16 years) Study to Explore Early Development – Teen (2017-2021) cross-sectional analyses: associations between adolescent predictors and adolescent SRS-2 score.

| Developmental, Behavioral, and Psychiatric Conditions | | | | | | | | |
| --- | --- | --- | --- | --- | --- | --- | --- | --- |
|  | POP | |  | ASD | |  | DD | |
| Predictors | Estimate | p-value |  | Estimate | p-value |  | Estimate | p-value |
| ADHD | 7.60 | <0.001 |  | 5.70 | 0.004 |  | 8.68 | <0.001 |
| Anxiety | 5.66 | <0.001 |  | 3.87 | 0.063 |  | 8.05 | <0.001 |
| Behavioral or Conduct Problems | 10.31 | 0.001 |  | 11.10 | <0.001 |  | 10.76 | <0.001 |
| Depression | 8.01 | <0.001 |  | 0.06 | 0.986 |  | 10.35 | <0.001 |
| Developmental Concerns | 13.78 | <0.001 |  | 11.29 | <0.001 |  | 12.28 | <0.001 |
| Externalizing | 7.32 | <0.001 |  | 8.57 | <0.001 |  | 9.33 | <0.001 |
| Internalizing | 6.55 | <0.001 |  | 4.11 | 0.045 |  | 8.51 | <0.001 |
| Learning Disability | 6.91 | <0.001 |  | 5.40 | 0.006 |  | 9.36 | <0.001 |
| Movement or Motor Problems | 2.58 | 0.640 |  | 8.78 | 0.001 |  | 5.71 | 0.027 |
| Sensory Integration Disorder | 3.42 | 0.716 |  | 8.47 | <0.001 |  | 8.57 | 0.002 |
| Speech or Language Disorder | 10.92 | <0.001 |  | 10.01 | <0.001 |  | 8.60 | <0.001 |
| Other Conditions | | | | | | | | |
| Allergy/Eczema | 1.11 | 0.270 |  | 0.84 | 0.716 |  | 1.56 | 0.312 |
| Asthma | -0.23 | 0.896 |  | 0.76 | 0.827 |  | -1.12 | 0.640 |
| GIS | 2.94 | 0.050 |  | 5.25 | 0.039 |  | 5.34 | 0.004 |
| CSHQ Parasomnia Score | 0.82 | 0.228 |  | 2.74 | 0.002 |  | 3.65 | <0.001 |
| Epilepsy/Seizures | 14.30 | 0.082 |  | 7.94 | 0.043 |  | 1.44 | 0.804 |
| Self-injurious Behavior | 3.36 | 0.537 |  | 9.57 | 0.001 |  | 10.65 | 0.033 |
| CSHQ Sleep Duration Score | 1.27 | 0.011 |  | 3.50 | <0.001 |  | 2.30 | 0.002 |

Models included group, study site, child sex, child age at the Mullen assessment, maternal age, maternal education, and maternal percent poverty level as covariates (continuous covariates are centered and scaled across diagnostic groups). Analysis of Variance models were used to test for the interaction between group and each predictor while controlling for covariates. Test statistics for seizure are not reported for the POP group because only one adolescent in the POP group reported the presence of seizure. POP=population controls; ASD=autism spectrum disorder; DD=developmental delay; ADHD=attention deficit hyperactivity disorder; GIS=gastrointestinal symptoms. Effective sample size varied slightly per model because incomplete cases were dropped on a per-model basis, but the effective sample sizes did not differ by more than 7% within any model group. Dashes indicate that the sample size was too small to allow for model fitting. Estimates are regression coefficients.

Supplemental Table 5. Longitudinal analyses: Associations between Study to Explore Early Development – Phase 1 (2007-2011) early childhood predictors (ages 2-5) and SRS-2 score change from childhood to adolescence (ages 12-16) in the Study to Explore Early Development – Teen (2017-2021).

Supplemental Table 5. Longitudinal analyses: associations between childhood predictors and adolescent SRS-2 scores.

| Developmental, Behavioral, and Psychiatric Conditions | | | | | | | | |
| --- | --- | --- | --- | --- | --- | --- | --- | --- |
|  | POP | |  | ASD | |  | DD | |
| Predictors | Estimate | p-value |  | Estimate | p-value |  | Estimate | p-value |
| ADHD: Yes | -1.12 | 0.980 |  | -1.43 | 0.836 |  | 1.44 | 0.811 |
| CBCL Externalizing T-score | 0.87 | 0.594 |  | 0.54 | 0.811 |  | 1.53 | 0.474 |
| CBCL Internalizing T-score | 0.27 | 0.836 |  | -0.26 | 0.972 |  | 0.46 | 0.811 |
| Learning Disability: Yes | 19.02 | 0.043 |  | 1.58 | 0.811 |  | 1.20 | 0.811 |
| Movement or Motor Problems: Yes | 5.42 | 0.508 |  | -2.55 | 0.677 |  | 1.60 | 0.797 |
| Mullen ELC Standard Score | -2.04 | 0.143 |  | -4.85 | <0.001 |  | -4.01 | <0.001 |
| Self-Injurious Behavior: Yes | - | - |  | -3.18 | 0.811 |  | 12.81 | 0.017 |
| Speech or Language Disorder: Yes | 1.95 | 0.602 |  | -0.05 | 0.994 |  | 1.21 | 0.797 |
| Other Conditions | | | | | | | | |
| Allergy/Eczema | 0.60 | 0.811 |  | -1.64 | 0.766 |  | -0.82 | 0.811 |
| Asthma: Yes | 2.54 | 0.357 |  | 1.64 | 0.811 |  | -2.46 | 0.603 |
| Birth Defects: Yes | 0.29 | 0.994 |  | 4.52 | 0.602 |  | 3.12 | 0.594 |
| GIS: Yes | 0.34 | 0.932 |  | 1.24 | 0.811 |  | 0.14 | 0.994 |
| Hearing Problems: Yes | -3.72 | 0.766 |  | 2.25 | 0.811 |  | 0.08 | 0.994 |
| Pneumonia: Yes | 3.05 | 0.508 |  | 0.96 | 0.932 |  | 2.80 | 0.603 |
| Respiratory Syncytial Virus: Yes | -0.05 | 0.994 |  | -1.95 | 0.811 |  | -1.17 | 0.811 |
| Seizure | - | - |  | 4.60 | 0.797 |  | 4.47 | 0.602 |
| Sensory Integration Disorder: Yes | - | - |  | 0.00 | 1.000 |  | -0.97 | 0.811 |
| CSHQ Sleep Total Score | -1.00 | 0.357 |  | 1.11 | 0.594 |  | 0.36 | 0.811 |

Note: Models included group, study site, child sex, child age at the Mullen assessment, maternal age, maternal education, and maternal percent poverty level as covariates (continuous covariates are centered and scaled across diagnostic groups). Analysis of Variance models were used to test for the interaction between group and each predictor while controlling for covariates. Test statistics for seizure, self-injurious behavior, and sensory integration disorder are not reported for the POP group due to low prevalence. CBCL= Child Behavior Checklist POP=population controls; ASD=autism spectrum disorder; DD=developmental delay; ADHD=attention deficit hyperactivity disorder; GIS=gastrointestinal symptoms. Effective sample size varied minimally per model because incomplete cases were dropped on a per-model basis, but the effective sample sizes did not differ by more than 5% within any model group. Dashes indicate that the sample size was too small to allow for model fitting. Estimates are regression coefficients.
